# Meningioma epigenetic grouping reveals biologic drivers and therapeutic vulnerabilities

**DOI:** 10.1101/2020.11.23.20237495

**Authors:** Abrar Choudhury, Stephen T. Magill, Charlotte D. Eaton, Briana C. Prager, William C. Chen, Kyounghee Seo, Calixto-Hope G. Lucas, Javier E. Villanueva-Meyer, Tai-Chung Lam, Jenny Kan-Suen Pu, Lai-Fung Li, Gilberto Ka-Kit Leung, Harish N. Vasudevan, S. John Liu, Jason W. Chan, Zhixin Qiu, Michael Y. Zhang, Michael V. Martin, Matthew S. Susko, Steve E. Braunstein, Nancy Ann Oberheim Bush, Jessica Schulte, Nicholas Butowski, Penny K. Sneed, Mitchel S. Berger, Arie Perry, Joanna J. Phillips, David A. Solomon, Joseph F. Costello, Michael W. McDermott, Jeremy N. Rich, David R. Raleigh

**Affiliations:** Department of Radiation Oncology, University of California San Francisco, San Francisco, CA 94143, USA; Department of Neurological Surgery, University of California San Francisco, San Francisco, CA 94143, USA; Medical Scientist Training Program, University of California San Francisco, San Francisco, CA 94143, USA; Biomedical Sciences Graduate Program, University of California, San Francisco, San Francisco, CA 94143, USA; Department of Medicine, University of California San Diego, San Diego, CA 92093, USA; Department of Pathology, University of California San Francisco, San Francisco, CA 94143, USA; Department of Radiology and Biomedical Imaging, University of California San Francisco, San Francisco, CA 94143, USA; Department of Clinical Oncology, The University of Hong Kong, Pokfulam, Hong Kong; Department of Neurosurgery, Division of Neurosurgery, The University of Hong Kong, Pokfulam, Hong Kong; Department of Neurology, University of California San Francisco, San Francisco, CA 94143, USA; Miami Neuroscience Institute, Baptist Health, Miami, FL 33176, USA

## Abstract

Meningiomas arising from the meningothelial central nervous system lining are the most common primary intracranial tumors, and a significant cause of neurologic morbidity and mortality^1^. There are no effective medical therapies for meningioma patients^2,3^, and new treatments have been encumbered by limited understanding of meningioma biology. DNA methylation profiling provides robust classification of central nervous system tumors^4^, and can elucidate targets for molecular therapy^5^. Here we use DNA methylation profiling on 565 meningiomas integrated with genetic, transcriptomic, biochemical, and single-cell approaches to show meningiomas are comprised of 3 epigenetic groups with distinct clinical outcomes and biological features informing new treatments for meningioma patients. Merlin-intact meningiomas (group A, 34%) have the best outcomes and are distinguished by a novel apoptotic tumor suppressor function of *NF2*/Merlin. Immune-enriched meningiomas (group B, 38%) have intermediate outcomes and are distinguished by immune cell infiltration, *HLA* expression, and lymphatic vessels. Hypermitotic meningiomas (group C, 28%) have the worst outcomes and are distinguished by convergent genetic mechanisms misactivating the cell cycle. Consistently, we find cell cycle inhibitors block meningioma growth in cell culture, organoids, xenografts, and patients. Our results establish a framework for understanding meningioma biology, and provide preclinical rationale for new therapies to treat meningioma patients.

## MAIN TEXT

### Meningiomas are comprised of 3 epigenetic groups

To identify biologic drivers and therapeutic vulnerabilities underlying meningioma, DNA methylation profiling was performed on 565 meningiomas from patients with comprehensive clinical follow-up who were treated at 2 independent institutions from 1991 to 2019. Consistent with typical meningioma outcomes, local freedom from recurrence (LFFR) and overall survival (OS) were worse with higher World Health Organization (WHO) grade, recurrent presentation, or subtotal resection (**Extended Data Fig. 1a**). Meningiomas were stratified by institution into a 200-sample discovery cohort from the University of California San Francisco (median follow-up 6.3 years), and a consecutive 365-sample validation cohort from The University of Hong Kong (median follow-up 5.3 years) (**Extended Data Table 1**). Prior DNA methylation studies have identified epigenetic groups of meningiomas with distinct clinical outcomes^6–9^, suggesting DNA methylation profiles may encode biologic information that could inform new treatments for meningioma patients. However, meningiomas that are resistant to current therapies have an abundance of genomic deletions and amplifications^9,10^, which can bias DNA methylation analyses^11^. Prior studies have not accounted for these confounding factors, and have not provided mechanistic or functional validation of meningioma groups. Thus, opportunities exist to build on foundational observations from initial meningioma DNA methylation studies, particularly through new bioinformatic techniques clarifying epigenetic groups in the context of copy number variation (CNV), mechanistic interrogation of epigenetic groups, and functional discovery of new therapies for meningioma patients.

Unsupervised hierarchical clustering of DNA methylation probes, controlled for artifacts from genomic deletions and amplifications using the SeSAMe preprocessing pipeline^11^, revealed 3 epigenetic groups of meningiomas in the discovery cohort (**Fig. 1a**). K-means consensus clustering validated 3 groups as the optimal number in the discovery and validation cohorts (**Fig. 1b and Extended Data Fig. 1b**). To assign meningiomas from the validation cohort into epigenetic groups, a multi-class support vector machine classifier was constructed that performed with 97.9% accuracy when classifying random samples from the discovery cohort (95% CI 89.2-99.9%, p<2.2×10^−16^). Kaplan-Meier analyses of the discovery and validation cohorts showed epigenetic groups were distinguished by differences in LFFR and OS (**Fig. 1c**), and correlated with prognostic clinical features of grade, sex, prior radiotherapy, and location (**Fig. 1d and Extended Data Fig. 2a**)^12^. Nevertheless, epigenetic groups were independently prognostic for outcomes on Kaplan-Meier analysis across WHO grades (**Extended Data Fig. 2b, c**), and on multivariable regression accounting for grade, extent of resection and other clinical features (**Extended Data Table 2**).

**Fig. 1.**
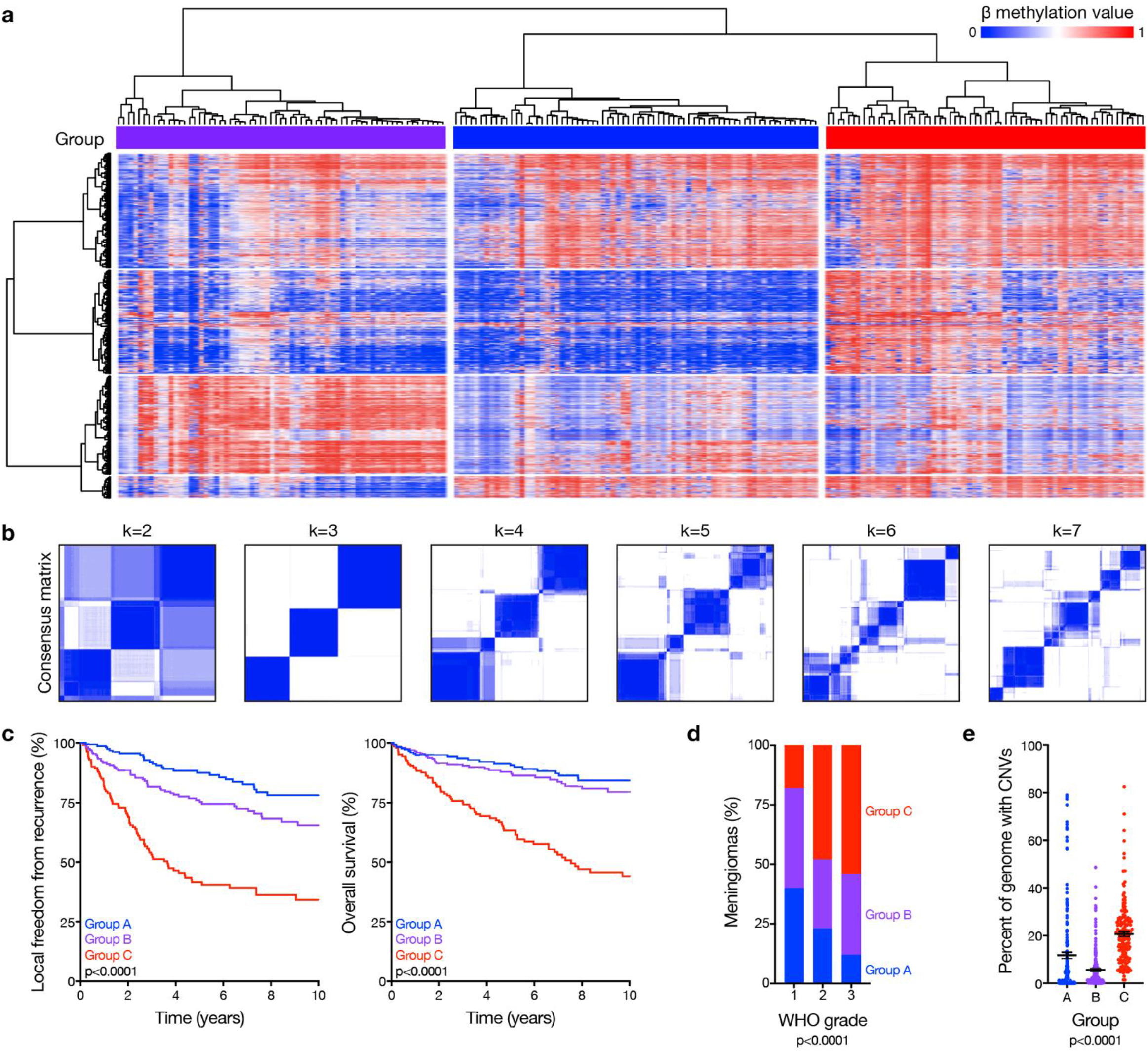
Meningioma is comprised of 3 epigenetic groups with distinct clinical outcomes. **a**, Unsupervised hierarchical clustering of meningiomas from the discovery cohort (n=200) using 2,000 differentially methylated DNA probes. **b**, K-means consensus clustering of meningiomas from the discovery and validation cohorts (n=565). **c**, Kaplan-Meier curves for meningioma local freedom from recurrence and overall survival (n=565) across epigenetic groups (Log-rank tests). **d**, Meningioma World Health Organization (WHO) grades (n=565) across epigenetic groups (Chi-squared test). **e**, Meningioma genomes (n=565) with copy number variations (CNVs) across epigenetic groups (ANOVA).

There were abundant CNVs across epigenetic groups (**Extended Data Fig. 3a**), underscoring the importance of accounting for genomic deletions and amplifications in meningioma DNA methylation profiles. Group C meningiomas had the worst outcomes (**Fig. 1c**) and had more CNVs compared to tumors from other groups (**Fig. 1e and Extended Data Fig. 3b**). Group A meningiomas had the best outcomes (**Fig. 1c**) and included 76% of the tumors with gain of chromosome 5 (**Extended Data Fig. 3b**), which is reported in meningiomas with favorable outcomes after surgery or radiotherapy^10,13^. Group B meningiomas had intermediate outcomes (**Fig. 1c**) and had fewer CNVs compared to tumors from other groups (**Fig. 1e**).

### Group A meningiomas are Merlin-intact

Meningiomas are common in patients with neurofibromatosis type 2, a complex autosomal disorder caused by loss of the tumor suppressor *NF2* on chromosome 22q^14^. *NF2* is also the most recurrently mutated gene in sporadic^15,16^ and radiation-induced^17^ meningiomas. Patients in the discovery or validation cohorts with neurofibromatosis type 2 (n=18) or radiation-induced meningiomas (n=34) overwhelmingly had group B or C meningiomas (96%), and *NF2* was lost in 86% of group B and C tumors, but only in 17% of group A meningiomas (**Fig. 2a**). RNA sequencing of the 200-sample discovery cohort confirmed lower *NF2* expression in group B and C meningiomas compared to group A tumors (**Fig. 2b**). Amplicon-based DNA sequencing on 65 samples from the discovery cohort confirmed that accounting for mutations, insertions, or deletions in *NF2* did not alter the frequency of *NF2* alterations in group A meningiomas (**Extended Data Table 3**). The combined distribution of *NF2* CNVs and somatic short variants showed 89% of group A meningiomas encoded at least 1 wildtype copy of *NF2* (**Fig. 2c**), which translated to expression of Merlin protein (**Fig. 2d**). *TRAF7* somatic short variants are mutually exclusive to *NF2* inactivating variants in meningiomas^15,18^, and were enriched in group A (Merlin-intact) meningiomas compared to tumors from other groups (**Extended Data Table 4**). However, many Merlin-intact meningiomas did not encode *TRAF7* somatic short variants, suggesting the epigenetic group of meningiomas with the best outcomes may not be unified by a single oncogenic mechanism. In support of this hypothesis, meningioma histologic subtypes associated with AKT1^E17K^ somatic short variants were also enriched in Merlin-intact meningiomas compared to tumors from other groups (**Extended Data Table 4**)^19^.

**Fig. 2.**
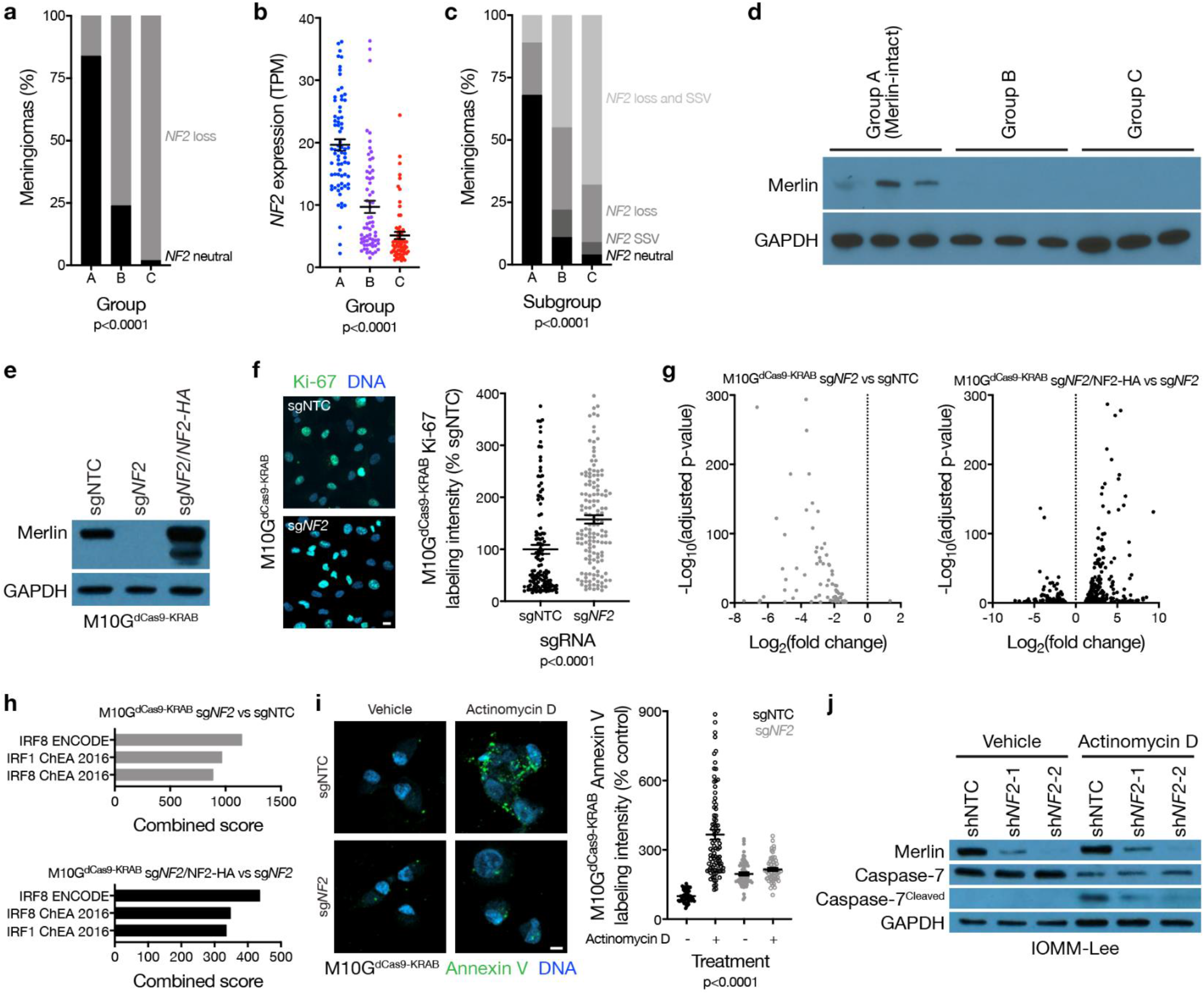
Meningioma loss of *NF2*/Merlin increases cell proliferation and decreases apoptosis. **a**, Meningioma DNA methylation-based analysis of copy number loss at the *NF2* locus (n=565) across epigenetic groups (Chi-squared test). **b**, Meningioma transcripts per million (TPM) expression of *NF2* (n=200) across epigenetic groups (ANOVA). **c**, Meningioma (n=65) *NF2* copy number loss and targeted sequencing of somatic short variants (SSVs) across epigenetic groups (Chi-squared test). **d**, Immunoblot for Merlin and GAPDH in 3 meningiomas with *NF2* loss at the *NF2* locus from each epigenetic group. **e**, Immunoblot for Merlin and GAPDH in M10G^dCas9-KRAB^ cells stably expressing a non-targeting control single-guide RNA (sgNTC), a single-guide RNA targeting *NF2* (sg*NF2*), or sg*NF2* with concurrent Merlin rescue using an exogenous *NF2* construct with an N-terminal HA tag. **f**, Confocal immunofluorescence microscopy and quantification of Ki-67 in M10G^dCas9-KRAB^ cells from **e** (Student’s t test). DNA is marked with Hoechst 33342. Scale bar 10 μM. **g**, Volcano plots of relative gene expression from RNA sequencing of M10G^dCas9-KRAB^ cells in **e. h**, Gene ontology analysis of differentially expressed genes downregulated from RNA sequencing of M10G^dCas9-KRAB^ cells in **e. i**, Confocal immunofluorescence microscopy and quantification of Annexin V in M10G^dCas9-KRAB^ cells from **e** treated with actinomycin D 0.5 μg/m or vehicle control for 48 hours (ANOVA). DNA is marked with DAPI. Scale bar 10 μM. **j**, Immunoblot for Merlin, Caspase-7, cleaved Caspase-7, or GAPDH in IOMM-Lee cells stably expressing non-targeting control shRNAs (shNTC) or shRNA targeting *NF2* (sh*NF2*-1 and sh*NF2*-2) treated with actinomycin D or vehicle control as in **i**.

Merlin has pleiotropic tumor suppressor functions at the plasma membrane^20,21^ and nucleus^22^ in schwannoma cells, but Merlin tumor suppressor functions in meningiomas are incompletely understood. M10G and IOMM-Lee meningioma cells express Merlin^23,24^, and Merlin suppression in these cells increased meningioma cell proliferation (**Fig. 2e, f and Extended Data Fig. 4a, b, c**). To identify gene expression programs underlying Merlin tumor suppressor functions in meningiomas, RNA sequencing was performed on triplicate M10G cultures stably expressing the CRISPR interference (CRISPRi) components dCas9-KRAB^25^ and non-targeting control sgRNA (sgNTC), sgRNA targeting *NF2* (sg*NF2*), or sg*NF2* with *NF2* rescue (**Fig. 2e**). Differential expression and ontology analyses revealed Merlin induced apoptotic interferon pathways^26–29^ previously unknown to be associated with either Merlin tumor suppressor functions or meningioma biology (**Fig. 2g, h and Extended Data Table 5**). To test this mechanistically, M10G^dCas9-KRAB^ and IOMM-Lee cultures were treated with the cytotoxic chemotherapy actinomycin D, revealing Merlin protected meningioma cells from apoptosis (**Fig. 2i, j, and Extended Data Fig. 4d**). These data shed light on a novel apoptotic tumor suppressor function of Merlin underlying the epigenetic group of meningiomas with the best outcomes that is lost in groups that are resistant to current therapies (**Fig. 1c**).

### Group B meningiomas are Immune-enriched

The relative reduction in group B meningioma CNVs suggests bulk bioinformatic analyses of meningiomas may be influenced by non-tumor cells in the meningioma microenvironment (**Fig. 1e**). SeSAMe^11^ cell-type deconvolution of DNA methylation profiles revealed enrichment of immune cells in group B meningiomas compared to tumors from other groups (**Fig. 3a**). PAMES^30^ tumor purity analysis and xCell^31^ RNA sequencing deconvolution validated these findings (**Fig. 3b, c, and Extended Data Fig. 5a,b**). Differential expression and gene ontology analyses showed enrichment of immune genes in Immune-enriched (group B) meningiomas (**Extended Data Fig. 5c and Extended Data Table 6**), and immunohistochemistry revealed T cell enrichment in Immune-enriched meningiomas compared to tumors from other groups (**Fig. 3d**). To validate these findings, single-cell RNA sequencing was performed on 57,114 cells from 8 meningioma samples representing each epigenetic group (**Extended Data Fig. 6a**). Reduced dimensionality clusters of meningioma and non-meningioma cells were identified by chromosome 22q loss (**Extended Data Fig. 6b**). Non-meningioma cell clusters with retained chromosome 22q were classified by expression of immune, neural, and vascular marker genes (**Extended Data Fig. 6c, d and Extended Data Table 7**). Meningioma cell clusters with chromosome 22q loss were distinguished by uniquely expressed marker genes (**Extended Data Fig. 6c, d and Extended Data Table 7**). In sum, single-cell transcriptomes revealed more immune cells in Immune-enriched meningiomas compared to tumors from other groups (**Fig. 3e**). Moreover, analysis of our previously reported DNA methylation profiles on 86 spatially distinct meningioma samples from 13 tumors^23^ revealed 92% of samples classified in concordance with the consensus epigenetic group of each tumor (**Extended Data Fig. 5d**). Thus, epigenetic grouping of meningiomas is not confounded by intratumor heterogeneity or sampling bias, suggesting coordinated genetic mechanisms underlie a novel group of Immune-enriched meningiomas that is distinguished by immune cell infiltration.

**Fig. 3.**
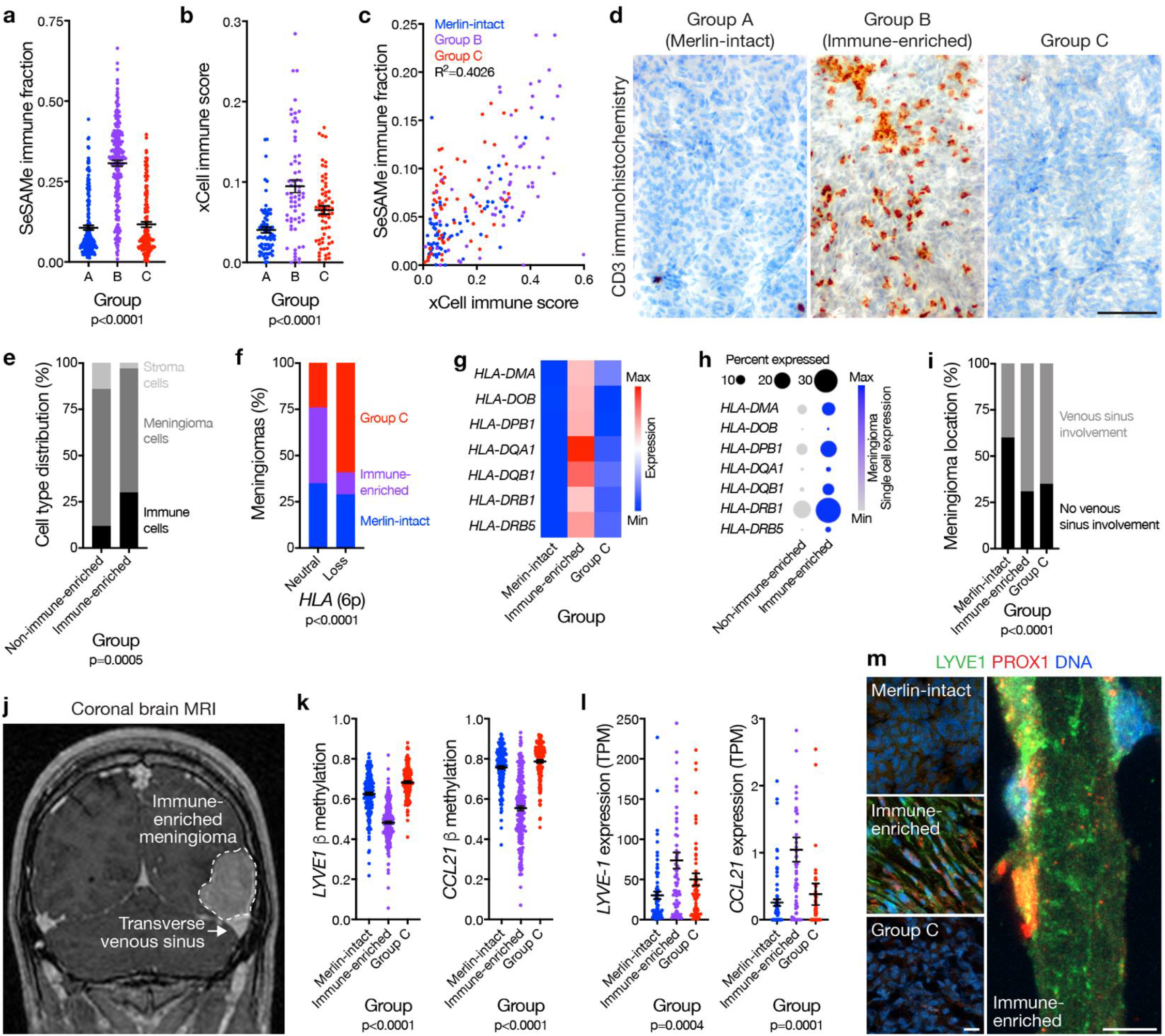
Meningioma immune enrichment is associated with HLA expression and meningeal lymphatics. **a**, Meningioma DNA methylation-based estimation of leukocyte fraction (n=565) across epigenetic groups (ANOVA). **b**, Meningioma RNA sequencing-based xCell immune score (n=200) across epigenetic groups (ANOVA). **c**, Correlation of meningioma leukocyte fraction from **a** and immune score from **b** (n=200) across epigenetic groups. **d**, Representative images of meningioma CD3 immunohistochemistry (n=87) across epigenetic groups (p<0.0001, Chi-squared test). Scale bar 100 μM. **e**, Single-cell RNA sequencing-based quantification of immune, stroma, and meningioma cells in Immune-enriched (n=5) and non-Immune-enriched (n=3) meningioma samples (Chi-squared test). **f**, Meningioma DNA methylation-based analysis of copy number loss at the *HLA* locus (n=565) across epigenetic groups (Chi-squared test). **g**, Meningioma RNA sequencing relative expression of *HLA* genes (n=200) across epigenetic group. **h**, Meningioma single-cell RNA sequencing relative expression of *HLA* genes across Immune-enriched (n=5) and non-Immune-enriched (n=3) meningioma samples. Circle size denotes percentage of cells. Circle shading denotes average expression. **i, j** Meningioma location on preoperative magnetic resonance imaging (n=169) across epigenetic groups (Chi-squared test). Representative image shown. **k**, Meningioma DNA methylation (n=565) of *LYVE-1* (cg26455970) or *CCL21* (cg27443224) across epigenetic groups (ANOVA). **l**, Meningioma transcripts per million (TPM) expression (n=200) of *LYVE-1* or *CCL21* across epigenetic groups (ANOVA). **m**, Representative images of meningioma LYVE1 and PROX1 confocal immunofluorescence microscopy across epigenetic groups (n=12). DNA is marked with Hoechst 33342. Scale bars 10 μM.

*HLA* loss can decrease immune infiltration in cancer^32^. 88% of meningiomas with *HLA* loss were Merlin-intact or group C tumors (**Fig. 3f**), and *HLA* expression was increased in Immune-enriched meningiomas compared to tumors from other groups (**Fig. 3g**). Single-cell transcriptomes showed increased *HLA* expression in Immune-enriched meningioma cells compared to meningioma cells from other groups (**Fig. 3h**), suggesting *HLA* differences across epigenetic groups were not confounded by non-tumor cells. Analysis of our previously reported matched whole exome sequencing and DNA methylation profiling on 25 meningiomas overlapping with the discovery cohort revealed no instances of *HLA* loss of heterozygosity in Immune-enriched meningiomas (9 Immune-enriched, 16 non-Immune-enriched)^9^. Thus, *HLA* expression may contribute to immune infiltration in Immune-enriched meningiomas compared to tumors from other groups.

Meningeal lymphatic vessels are necessary for intracranial immune surveillance^33–36^, and are concentrated near dural venous sinuses^33^, but are not known to influence intracranial tumor immune infiltration. Preoperative magnetic resonance imaging from the discovery cohort showed both Immune-enriched and group C meningiomas were more likely to involve dural venous sinuses than Merlin-intact tumors (**Fig. 3i, j**), suggesting immune infiltration and meningeal lymphatics in Immune-enriched meningiomas were not solely a byproduct of tumor location. Immune-enriched meningiomas had hypomethylation (**Fig. 3k**) and increased expression (**Fig. 3l**) of meningeal lymphatic genes such as *LYVE1* and *CCL21* compared to tumors from other groups^36–38^. Immunofluorescence for LYVE1 and the lymphatic marker PROX1^39^ confirmed lymphatic enrichment in Immune-enriched meningiomas compared to tumors from other groups (**Fig. 3m**). Thus, in addition to *HLA* expression, lymphatic vessels may also contribute to immune infiltration in Immune-enriched meningiomas compared to tumors from other groups.

### Group C meningiomas are Hypermitotic

High-grade meningiomas are defined by brisk cell proliferation leading to local recurrence and death in 50-90% of patients^40,41^. We previously reported FOXM1 is a key transcription factor for meningioma cell proliferation^9^, and FOXM1 target genes are reliable biomarkers for meningioma recurrence^42^. Cell proliferation was highest in group C meningiomas (**Fig. 4a**), and RNA sequencing and gene ontology analysis revealed the FOXM1 transcriptional program was enriched in Hypermitotic (group C) meningiomas compared to tumors from other groups (**Fig. 4b, Extended Data Fig. 7a, and Extended Data Table 6**). Immunohistochemistry and immunofluorescence showed FOXM1 protein localized to dividing meningioma cells, and correlated with meningioma cell proliferation across epigenetic groups (**Fig. 4c, d**). To define FOXM1 targets in meningioma, differentially expressed genes with FOXM1 binding motifs were analyzed in our previously reported matched RNA sequencing, H3K27ac ChIP sequencing, and DNA methylation profiling on 25 meningiomas (15 Hypermitotic, 10 non-Hypermitotic)^43^. FOXM1 target genes in Hypermitotic meningiomas regulated the DNA damage response, the cell cycle, and tumor metabolism (**Extended Data Fig. 7b**), suggesting FOXM1 promotes treatment resistance in the epigenetic group of meningiomas with the worst outcomes after treatment with current therapies (**Fig. 1c**).

**Fig. 4.**
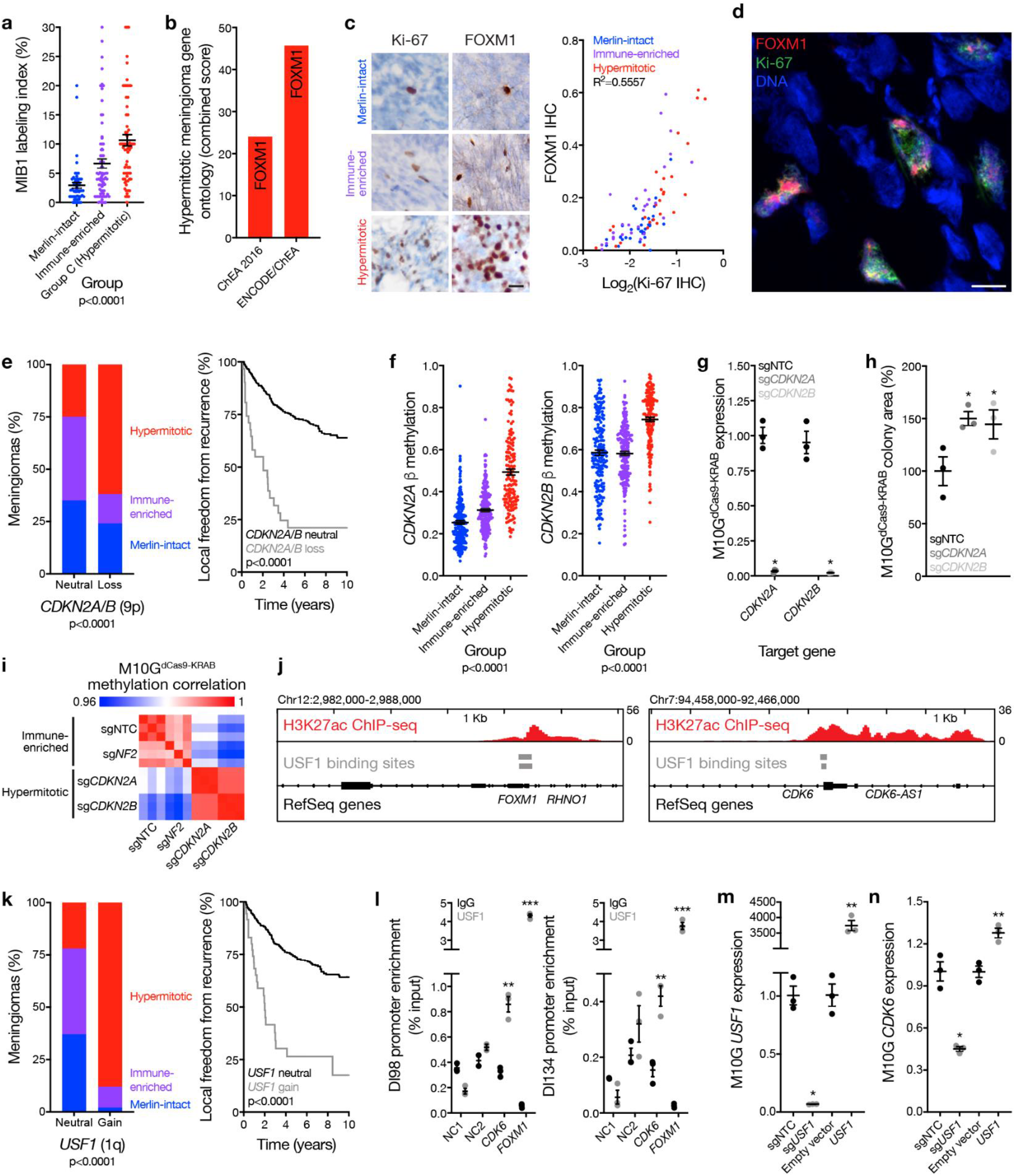
Convergent genetic mechanisms misactivate the cell cycle in meningioma. **a**, MIB1 labeling index from meningioma clinical pathology reports (n=206) across epigenetic groups (ANOVA). **b**, Gene ontology analysis of differentially expressed genes in Hypermitotic meningiomas compared to tumors from other groups. ChEA, ChIP-X Enrichment Analysis. **c**, Representative images of meningioma Ki-67 and FOXM1 immunohistochemistry and correlated quantification (n=92) across meningioma groups. Scale bar 10 μM. **d**, Representative image of meningioma Ki-67 and FOXM1 confocal immunofluorescence microscopy. DNA is marked with DAPI. Scale bar 10 μM. **e**, Meningioma DNA methylation-based analysis of copy number loss at the *CDKN2A/B* locus across epigenetic groups (left, n=565, Chi-squared test), and Kaplan-Meier curve for meningioma local freedom from recurrence stratified by *CDKN2A/B* copy number status (right, n=565, Log-rank test). **f**, Meningioma DNA methylation (n=565) of *CDKN2A* (cg26349275) or *CDKN2B* (cg208390209) across epigenetic groups (ANOVA). **g**, QPCR for *CDKN2A* or *CDKN2B* in M10G^dCas9-KRAB^ cells expressing a non-targeting control single-guide RNA (sgNTC), a single-guide RNA targeting the p16^INK4A^ isoform of *CDKN2A* (sg*CDKN2A*), or a single-guide RNA targeting *CDKN2B* (sg*CDKN2B*) (*p<0.05, Student’s t test). **h**, Relative colony area of M10G^dCas9-KRAB^ cells expressing sgNTC, sg*CDKN2A*, or sg*CDKN2B* after 10 days of clonogenic growth (*p<0.05, Student’s t test). **i**, Pearson correlation heatmap of all methylation probes for M10G^dCas9-KRAB^ cells expressing sgNTC, sg*NF2*, sg*CDKN2A*, or sg*CDKN2B* (n=3 per condition). **j**, H3K27ac ChIP sequencing tracks comparing *CDK6* and *FOXM1* loci across meningiomas (n=25) to normal neural cell and tissue samples (ChIP Atlas). USF1 binding sites are annotated. **k**, Meningioma DNA methylation-based analysis of copy number loss at the *USF1* locus across epigenetic groups (left, n=565, Chi-squared test), and Kaplan-Meier curve for meningioma local freedom from recurrence stratified by *USF1* copy number status (right, n=565, Log-rank test). **l**, ChIP-QPCR after USF1 pulldown in DI98 and DI134 meningioma cells for the *CDK6* or *FOXM1* promoters (Student’s t test) compared to negative control primers targeting a gene desert (NC1) or a gene not predicted to be bound by USF1 (NC2) from ChIP sequencing. **m**, QPCR for *USF1* in M10G^dCas9-KRAB^ cells expressing sgNTC or a single-guide RNA targeting *USF1* (sg*USF1*); or M10G cells expressing an empty vector or a vector containing exogenous *USF1* (Student’s t test). **n**, QPCR for *CDK6* in M10G cells from **m** (Student’s t test). *p≤0.05, **p≤0.01, ***p≤0.0001.

Druggable somatic short variants in meningiomas are rare and are not associated with adverse outcomes^10,14–19,44–49^, with rare exceptions^50–53^. Differential expression of enhancers and super-enhancers within the consensus meningioma peakset revealed the enhancer landscape of Hypermitotic meningiomas was dominated by epigenetic mechanisms and transcription factors, such as FOXM1, that are impractical pharmacologic targets (**Extended Data Fig. 8**). However, Hypermitotic meningiomas had more large genomic deletions and amplifications compared to tumors from other groups (**Fig. 1e and Extended Data Fig. 3**). Thus, CNVs contributing to cell cycle misactivation may harbor therapeutic vulnerabilities that could be used to inform new treatments for meningioma patients.

Loss of the endogenous CDK4/6 inhibitors *CDKN2A/B* is associated with worse outcomes in multiple brain tumors^54^, including meningiomas^53^. 62% *CDKN2A/B* losses occurred in Hypermitotic meningiomas and were associated with worse LFFR (**Fig. 4e**). *CDKN2A*/B was also hypermethylated in Hypermitotic meningiomas compared to tumors from other groups (**Fig. 4f**), a reported alternate mechanism of CDK4/6 misactivation in cancer^55,56^. To identify reagents to study meningioma epigenetic groups, DNA methylation profiling and epigenetic classification were performed on 9 meningioma cell lines. M10G cells classified among Immune-enriched meningiomas and expressed *CDKN2A/B* (**Extended Data Table 5**). Stable suppression of *CDKN2A* or *CDKN2B* in M10G^dCas9-KRAB^ cells using sgRNAs increased cell proliferation (**Fig. 4g, h**), and re-classified triplicate M10G^dCas9-KRAB^ DNA methylation profiles among Hypermitotic meningiomas (**Fig. 4i**). In contrast, DNA methylation profiles of triplicate M10G^dCas9-KRAB^ cultures stably expressing sg*NF2* (**Fig. 2e**), which also increased cell proliferation (**Fig. 2f**), remained classified among Immune-enriched meningiomas (**Fig. 4i**). These data suggest coordinated genetic events underlie a novel group of Hypermitotic meningiomas that is distinguished by cell cycle misactivation.

Not all Hypermitotic meningiomas have *CDKN2A/B* loss (**Fig. 4e**). Thus, additional mechanisms must exist to drive the cell cycle in Hypermitotic tumors. Recurrent transcription factor binding motifs were present in H3K27ac ChIP sequencing troughs at the *CDK6* and *FOXM1* loci (**Extended Data Table 8**). *USF1* is a transcription factor on chromosome 1q, which is associated with poor outcomes in other brain tumors^57^. USF1 binding sites were occupied in the promoters of *CDK6* and *FOXM1* (**Fig. 4j**); 88% of *USF1* gains occurred in Hypermitotic meningiomas (**Fig. 4k and Extended Data Fig. 3b**); and *USF1* gain was associated with worse LFFR (**Fig. 4k**). Moreover, USF1 bound to the *CDK6* and *FOXM1* promoters (**Fig. 4I**) and induced *CDK6* expression (**Fig. 4m, n**) in meningioma cells, demonstrating USF1 is a regulator of *CDK6* and *FOXM1* in meningioma. In sum, convergent genetic mechanisms misactivate the cell cycle in Hypermitotic meningiomas, suggesting cytostatic therapies targeting the cell cycle may be effective treatments for meningiomas that are resistant to current therapies.

### Cell cycle inhibition blocks meningioma growth

In comparison to Merlin-intact meningiomas, Immune-enriched and Hypermitotic tumors (i) have intermediate and poor outcomes, respectively (**Fig. 1c, 5a**); (ii) exist along a continuum of cell cycle misactivation (**Fig. 4a**); and (iii) are distinguished by multiple resistance mechanisms diminishing the efficacy of cytotoxic therapies (**Fig. 2i, j, and Extended Data Fig. 7b**). Thus, to interrogate the cell cycle as a therapeutic target in meningioma, the clinical CDK4/6 inhibitors abemaciclib, palbociclib, and ribociclib^58^ were tested in cell culture, organoids, and xenografts using Immune-enriched and Hypermitotic meningioma cells (**Fig. 5a**). CDK4/6 inhibitors blocked clonogenic growth of Immune-enriched and Hypermitotic meningioma cells (**Fig. 5b**). Suppression of *CDKN2A* or *CDKN2B* increased the efficacy of CDK4/6 inhibition in M10G^dCas9-KRAB^ cells (**Fig. 5c**), but was not necessary for responses to treatment. To test this therapeutic strategy in the context of a tumor microenvironment, Hypermitotic meningioma cells were co-cultured with cerebral organoids comprised of pre-differentiated human pluripotent stem-cell derived astrocytes. We previously reported this model restores intratumor heterogeneity in meningioma cells^23^, and intratumor heterogeneity is a source of resistance to cancer treatments^59^. Nevertheless, CDK4/6 inhibition attenuated the growth of Hypermitotic meningioma cells in co-culture with cerebral organoids (**Fig. 5d**). To define the pharmacodynamics and efficacy of cell cycle inhibitors *in vivo*, RB-intact meningioma xenografts were treated with CDK4/6 inhibitors. CDK4/6 blockade decreased expression of phospho-RB (**Fig. 5e**), inhibited cell proliferation (**Fig. 5f**), attenuated xenograft growth (**Fig. 5g**), and prolonged survival in mice (**Fig. 5h**). These data provide preclinical rationale to treat patients with Immune-enriched or Hypermitotic meningiomas with cell cycle inhibitors. More broadly, our mechanistic and functional results validate the first biomarker-based molecular treatment for meningiomas with adverse outcomes.

**Figure 5.**
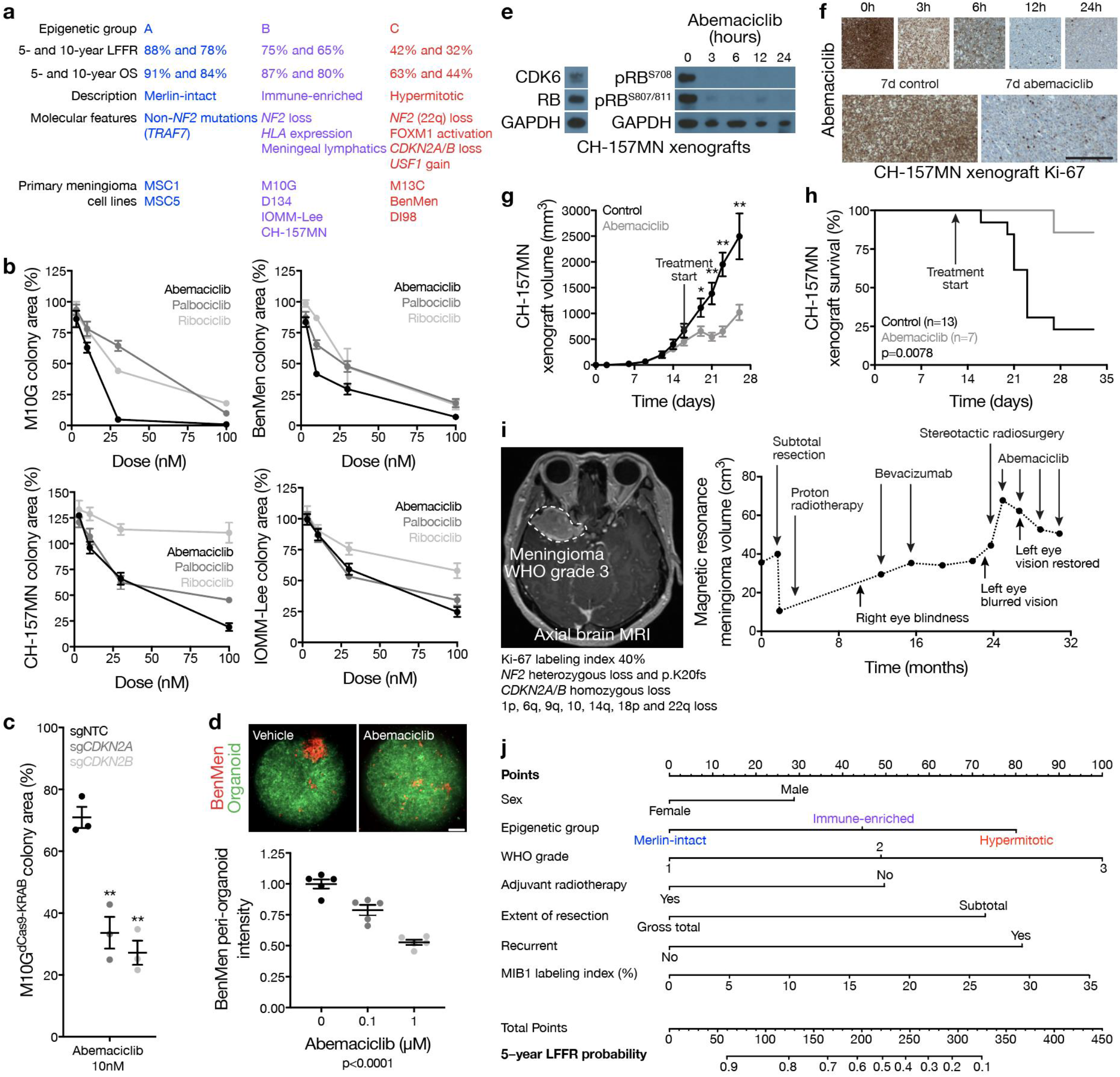
Cell cycle inhibition blocks meningioma growth. **a**, Clinical outcomes and molecular features of meningioma epigenetic groups, and cell lines classifying in each group. **b**, Relative colony area of M10G, BenMen, CH-157MN, or IOMM-Lee meningioma cells after 10 days of clonogenic growth and treatment with abemaciclib, ribociclib, or palbociclib. **c**, Relative colony area of M10G^dCas9-KRAB^ cells expressing sgNTC, sg*CDKN2A*, or sg*CDKN2B* after 10 days of clonogenic growth and treatment with abemaciclib (Student’s t test). Data are normalized to growth with vehicle treatment of each cell lines. **d**, Quantification of BenMen peri-organoid intensity after 10 days of growth treatment with abemaciclib or vehicle control (ANOVA). Representative image of meningioma (red) and organoid (green) cells is shown. Scale bars 100 μM. **e**, Representative immunoblots from subcutaneous CH-157MN xenograft tumors in NU/NU mice (left) harvested at intervals after a single treatment of abemaciclib (100 μg/g) via oral gavage (right). **f**, Representative images of CH-157MN xenograft Ki-67 immunohistochemistry after a single (top) or daily (bottom) treatment of abemaciclib (100 μg/g) via oral gavage. Scale bar 1 mM. **g**, Subcutaneous CH-157MN xenograft measurements in NU/NU mice during daily oral gavage with abemaciclib (100 μg/g) versus control (Student’s t test). **h**, Kaplan-Meier curve for subcutaneous CH-157MN xenograft overall survival in NU/NU mice (Log-rank test). **i**, Magnetic resonance imaging and molecular features of a representative meningioma (left) that was resistant to cytotoxic therapies but responded to cell cycle inhibition (right). **j**, Nomogram for meningioma local freedom from recurrence (LFFR, n=201) integrating established clinical features and novel epigenetic groups. Each variable contributes points (top row) to the total score, which estimates the probably of 5-year LFFR (bottom 2 rows) (https://william-c-chen.shinyapps.io/RaleighLab_MethylationSubgroupNomogram/). *p≤0.05, **p≤0.01, ***p≤0.0001.

In support of our preclinical investigations, we have observed encouraging early results with compassionate use of CDK4/6 inhibitors to treat human meningiomas that are resistant to surgery and radiotherapy (**Fig. 5i**). Formalized clinical trials to establish the efficacy of this and other molecular therapies for meningiomas will require rigorous patient selection and biologic stratification. In anticipation, data from the discovery and validation cohorts were used to develop prognostic nomograms integrating clinical features predicting meningioma recurrence alongside epigenetic groups predicting meningioma biology (**Fig. 5j and Extended Data Fig. 9a**). Although DNA methylation profiling is a powerful tool for biologic discovery, clinical adoption of this technique has been hampered by a lack of medical indications. The data presented here establish an urgent need for clinical DNA methylation profiling to stratify meningioma patients for molecular treatments. In the interim, recursive partitioning analysis of CNV profiles can be used to approximate epigenetic groups of meningiomas and improve prognostic models for meningioma recurrence (**Extended Data Fig. 9b-f**).

In conclusion, our results shed light on biochemical, cellular, and genomic mechanisms underlying the most common primary intracranial tumor. By accounting for artifacts from CNVs and integrating epigenetic, genetic, transcriptomic, biochemical, and single-cell approaches, we identify 3 epigenetic groups of meningiomas with distinct clinical and biological features that can inform new treatments for meningioma patients.

## Data Availability

The data generated from the research project will be made as widely and freely available as possible while safeguarding the privacy of subjects and protecting confidentiality. As per the NIH Genomic Data Sharing Policy NOT-OD-14-124, the large-scale human genomic data and relevant associated data generated by this research will be submitted to an NIH-designated data repository no later than 6 months after initial data submission for publication, or at the time of acceptance of first publication, whichever occurs first.

## Acknowledgements

The authors thank Aaron Tward and Adam Abate for comments and reagents, Anny Shai and the staff of the UCSF Brain Tumor Center Biorepository and Pathology Core, Tomoko Ozawa and the staff of the UCSF Brain Tumor Center Preclinical Therapeutics Core, and Eric Chow and the staff of the UCSF Center for Advanced Technology. This study was supported by the UCSF Wolfe Meningioma Program Project and NIH grants F30CA246808 and T32GM007618 to A.C.; NIH grant P50CA097257 to J.J.P.; the UCSF Wolfe Meningioma Program Project and NIH grant F32CA213944 to S.T.M.; the UCSF Wolfe Meningioma Program Project to C.D.E., J.E.V-M., H.N.V., S.E.B., N.A.O.B., J.S., and N.B.; and the UCSF Physician Scientist Scholar Program, the UCSF Wolfe Meningioma Program Project, and NIH grant K08CA212279 to D.R.R.

