## Extended Figures and Methods for "Meningioma epigenetic grouping reveals biologic drivers and therapeutic vulnerabilities"

**Extended Data Fig. 1. Clinical outcomes and subgroups in discovery and validation cohorts.**

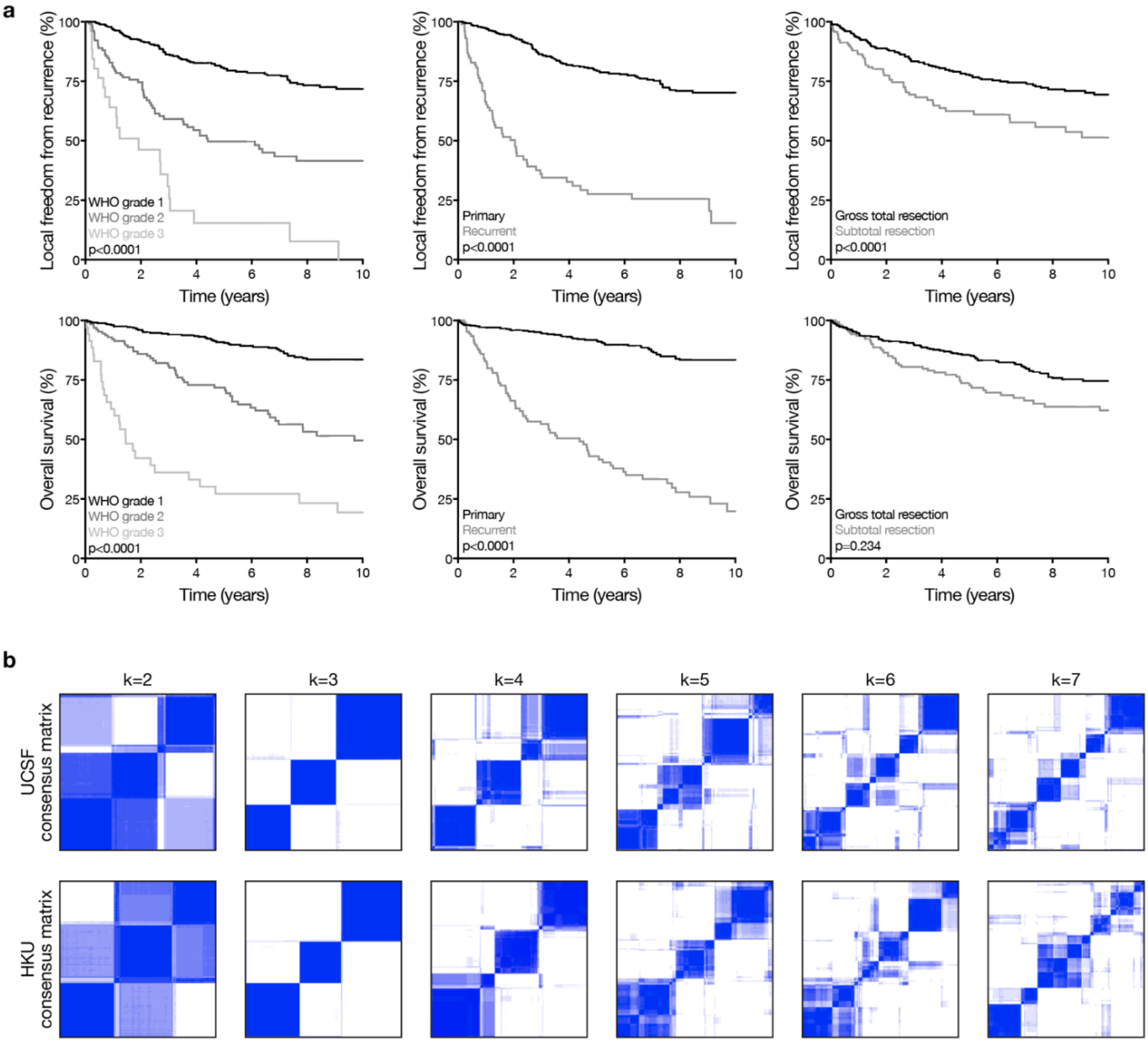

**Extended Data Fig. 1. Clinical outcomes and epigenetic groups in discovery and validation cohorts.**

**a**, Kaplan-Meier curves for meningioma local freedom from recurrence and overall survival (n=565) across clinical features (Log-rank tests). **b**, K-means consensus clustering of meningioma DNA methylation profiles from the discovery (n=200, UCSF) and validation (n=365, HKU) cohorts.

**Extended Data Fig. 2. Clinical correlations across meningioma groups.**

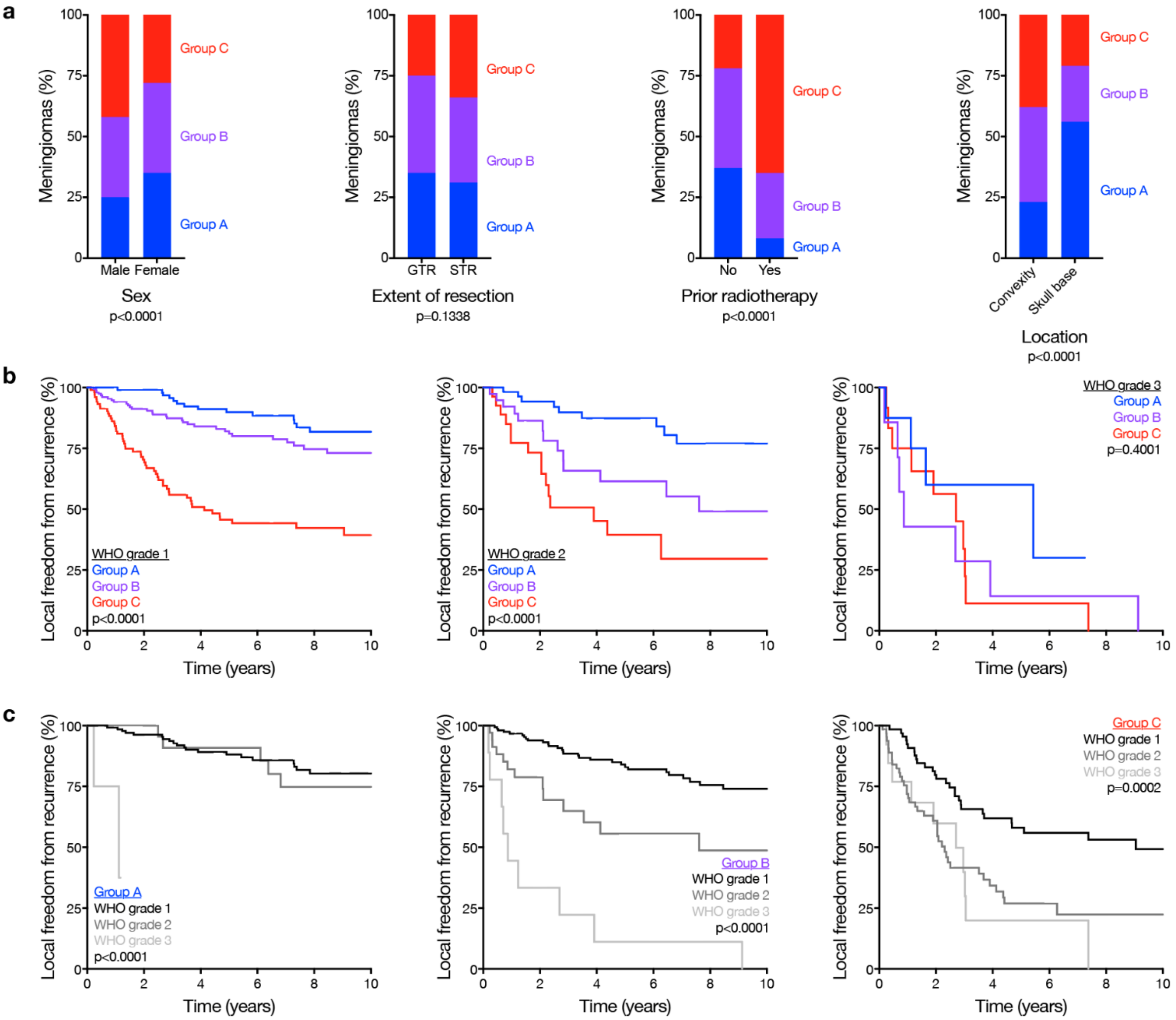

**Extended Data Fig. 2. Clinical correlations across meningioma groups.**

**a**, Meningioma clinical features (n=565) across epigenetic groups (Chi-squared tests). GTR, gross total resection. STR, subtotal resection. **b**, Kaplan-Meier curves for meningioma local freedom from recurrence (n=565) across WHO grades and epigenetic groups (Log-rank tests).

**Extended Data Fig. 3. Copy number variants across meningioma groups.**

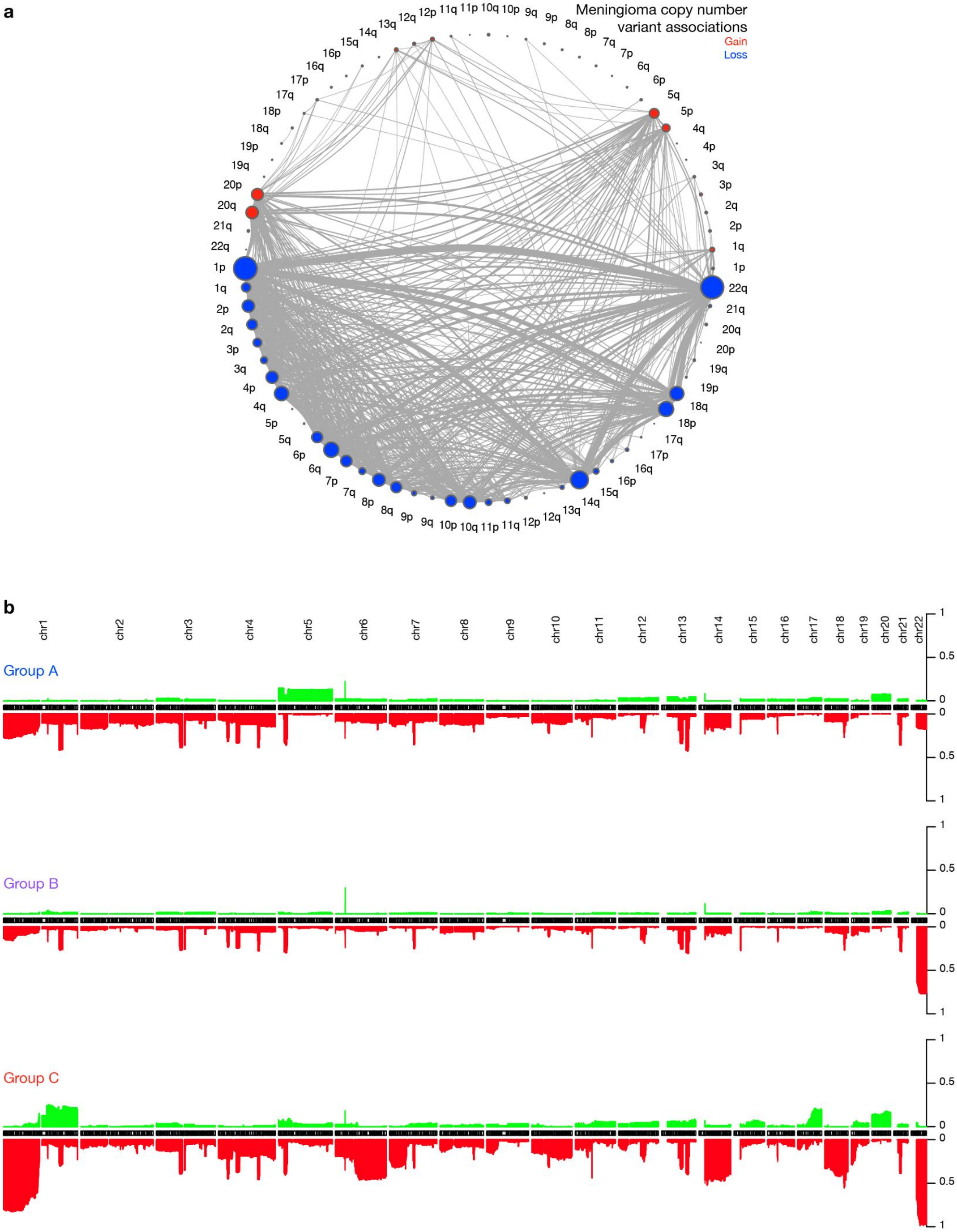

**Extended Data Fig. 3. Copy number variants across meningioma groups.**

**a**, Co-occurrence of chromosomal gains and losses in meningiomas (n=565). Circle size denotes frequency of copy number variants (CNVs). Line thickness denotes frequency of CNVs co-occurrence. **b**, Frequency of copy number losses (red) and gains (green) across meningioma epigenetic groups.

**Extended Data Fig. 4. Mechanisms of *NF2*/Merlin tumor suppression in meningioma.**

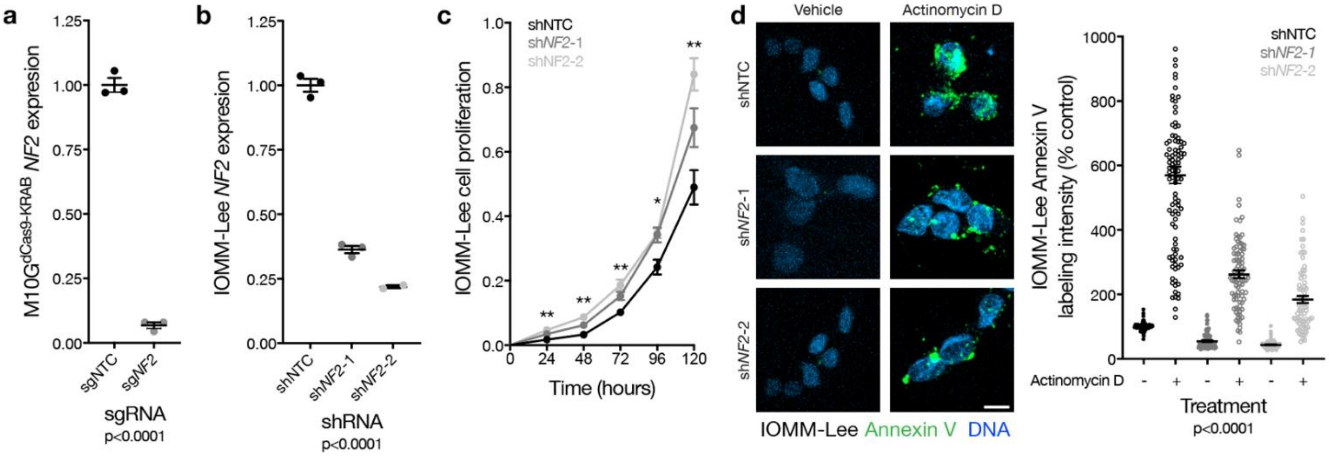

**Extended Data Fig. 4. Mechanisms of *NF2*/Merlin tumor suppression in meningioma.**

**a**, QPCR for *NF2* in M10G$^{dCas9\text{-}KRAB}$ cells expressing a non-targeting control single-guide RNA (sgNTC) or a single-guide RNA targeting *NF2* (sg*NF2*) (Student's t test). **b**, QPCR for *NF2* in IOMM-Lee cells expressing a non-targeting control shRNA (shNTC) or shRNAs targeting *NF2* (sh*NF2*-1 or sh*NF2*-2) (Student's t test). **c**, MTT cell proliferation of IOMM-Lee cells from **b**, normalized to shNTC at 24 hours (ANOVA). **d**, Confocal immunofluorescence microscopy and quantification of Annexin V in IOMM-Lee cells from **b** treated with actinomycin D 0.5µg/m or vehicle control (ANOVA). DNA is marked with DAPI. Scale bar 10 µM. *p≤0.05, **p≤0.01, ***p≤0.0001.

**Extended Data Fig. 5. Genomic and cellular characteristics of immune-enriched meningiomas.**

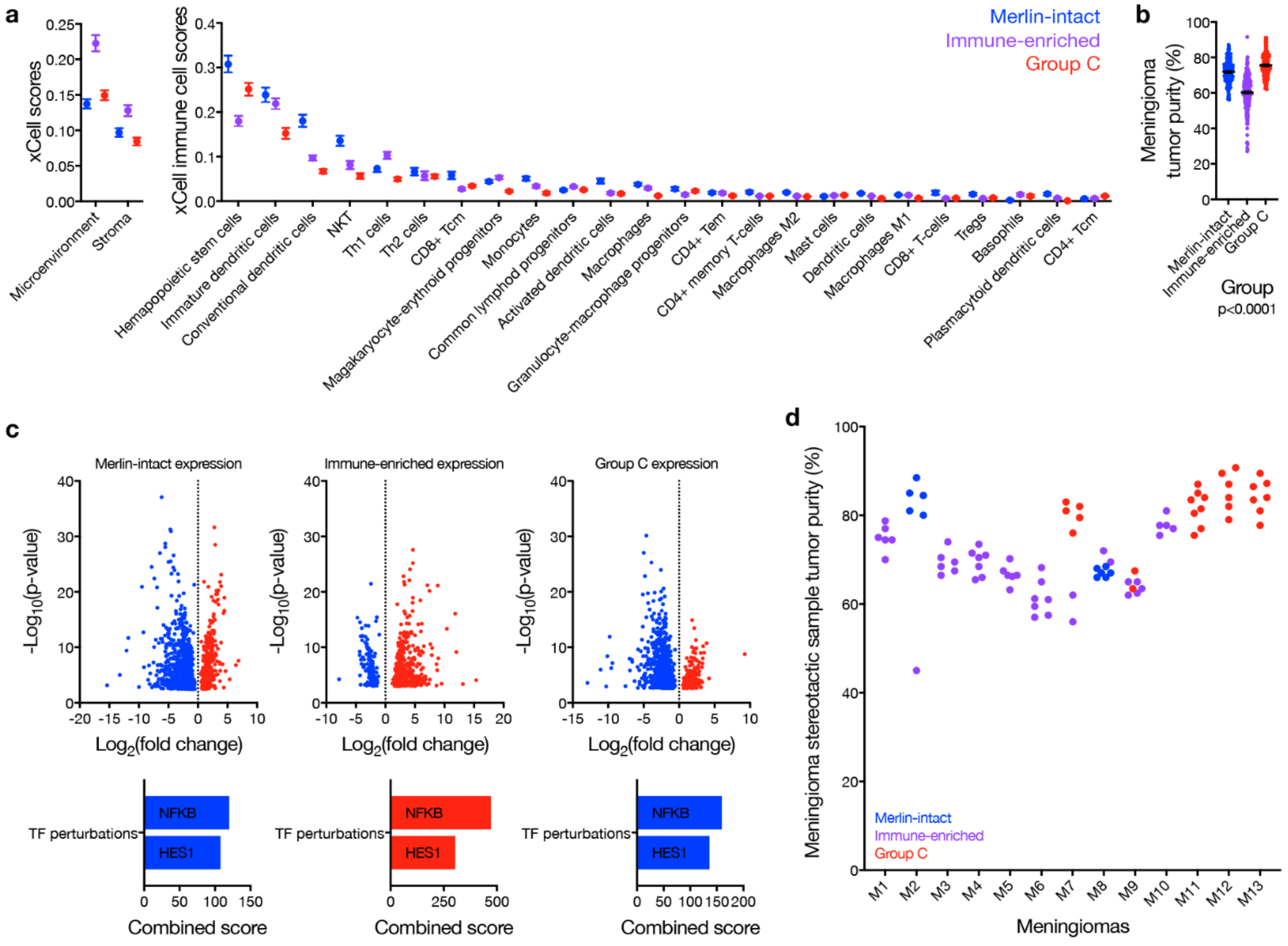

**Extended Data Fig. 5. Genomic and cellular characteristics of Immune-enriched meningiomas.**
**a**, Meningioma RNA sequencing xCell scores (n=200) across epigenetic groups for overall microenvironment and stroma (left), and individual immune cell types (right). **b,** Meningioma DNA methylation-based tumor purity (n=565) across epigenetic groups (ANOVA). **c**, Volcano plots of meningioma relative gene expression (n=200) across epigenetic groups, compared one group versus the others (top), and gene ontology transcription factor (TF) perturbation analysis (bottom), of differentially enriched (red) or suppressed (blue) genes. **d**, Meningioma DNA methylation-based tumor purity of 86 spatially-distinct samples from 13 meningiomas, colored by epigenetic group of each sample.

### Extended Data Fig. 6. Meningioma single cell RNA sequencing.

**a**

| Sample | Subgroup | Chr22q status | C0 | C1 | C2 | C3 | C4 | C5 | C6 | C7 | C8 | C9 | C10 | C11 | C12 | C13 | C14 | Total |
|---|---|---|---|---|---|---|---|---|---|---|---|---|---|---|---|---|---|---|
| MSC1 | Merlin-intact | Intact | 1441 | 191 | 11 | 152 | 17 | 46 | 28 | 6 | 5 | 578 | 19 | 29 | 11 | 3 | 1 | 2538 |
| MSC2 | Immune-enriched | Loss | 8 | 0 | 2 | 710 | 0 | 34 | 5 | 8 | 0 | 86 | 22 | 11 | 0 | 21 | 7 | 914 |
| MSC3 | Group C | Loss | 638 | 383 | 26 | 83 | 70 | 8 | 44 | 11 | 57 | 939 | 56 | 20 | 911 | 38 | 3 | 3287 |
| MSC4 | Merlin-intact | Intact | 855 | 1353 | 13 | 774 | 10 | 188 | 2170 | 1787 | 23 | 354 | 88 | 70 | 62 | 58 | 65 | 7870 |
| MSC4 dura | --- | Intact | 228 | 13 | 5 | 32 | 0 | 265 | 159 | 140 | 0 | 24 | 135 | 12 | 0 | 3 | 129 | 1145 |
| MSC5 | Immune-enriched | Loss | 6680 | 1510 | 18 | 1198 | 39 | 83 | 181 | 363 | 11 | 38 | 65 | 7 | 181 | 18 | 30 | 10422 |
| MSC5 brain/tumor interface | Immune-enriched | Loss | 198 | 6315 | 28 | 748 | 5 | 64 | 282 | 260 | 80 | 85 | 56 | 180 | 226 | 27 | 14 | 8568 |
| MSC5 dura | --- | Intact | 9 | 5 | 3 | 40 | 0 | 160 | 57 | 163 | 0 | 0 | 51 | 6 | 17 | 0 | 43 | 554 |
| MSC6 | Immune-enriched | Intact | 28 | 7 | 2509 | 330 | 1783 | 2546 | 265 | 154 | 1109 | 60 | 905 | 90 | 0 | 173 | 415 | 10374 |
| MSC6 brain/tumor interface | Immune-enriched | Intact | 35 | 19 | 3364 | 1164 | 2662 | 230 | 278 | 252 | 1847 | 264 | 554 | 1507 | 0 | 691 | 304 | 13171 |
| Total | --- | --- | 10120 | 9796 | 5979 | 5231 | 4586 | 3624 | 3469 | 3144 | 3132 | 2428 | 1951 | 1932 | 1408 | 1032 | 1011 | 58843 |

**b**

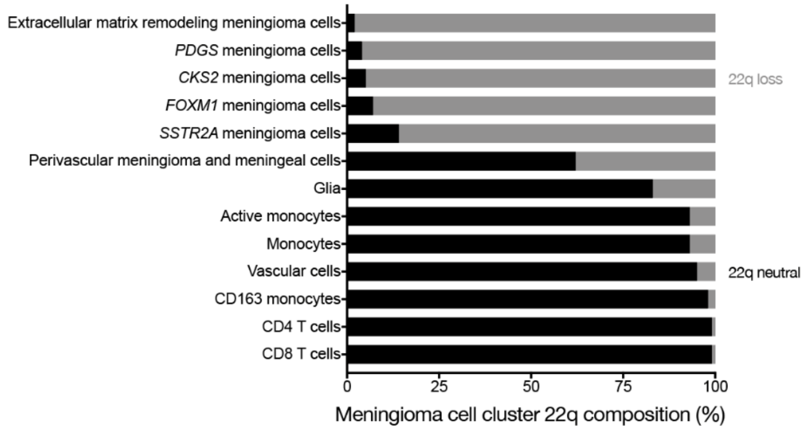


**c**

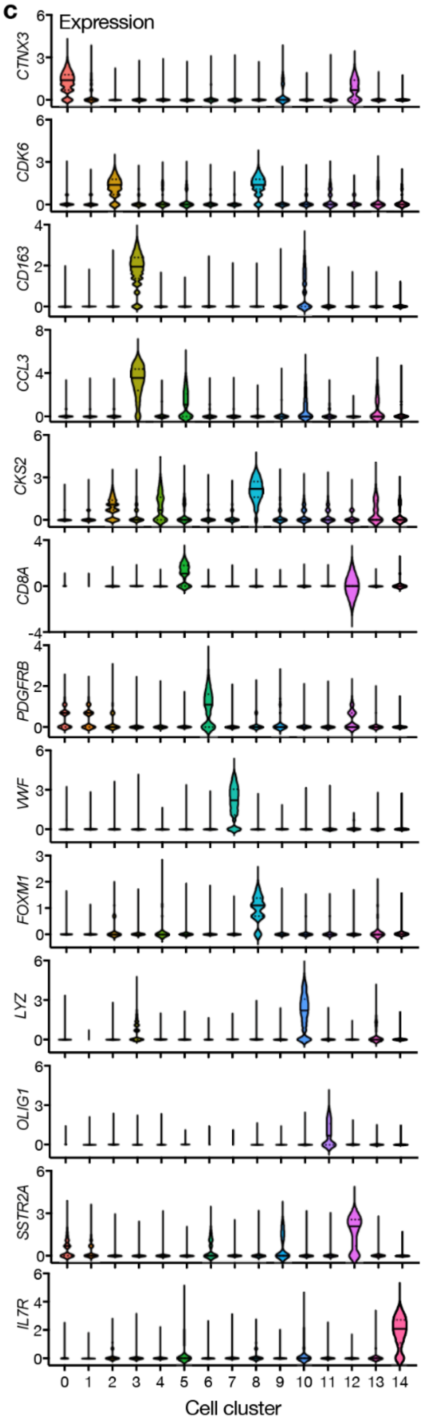


**d**

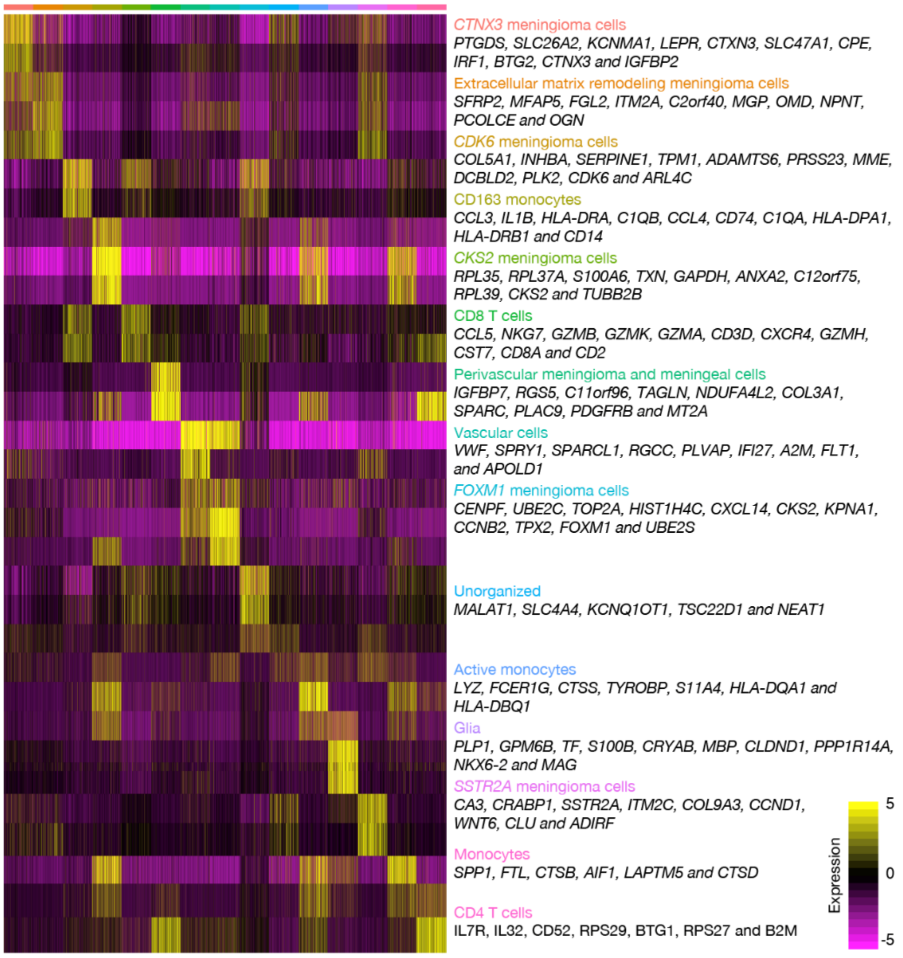

**Extended Data Fig. 6. Meningioma single-cell RNA sequencing.**

**a**, Cells in reduced dimensionality clusters from each sample analyzed using single-cell RNA sequencing. Epigenetic groups and chromosome 22q (Chr22q) status of meningioma samples are annotated. **b**, Percentage of cells with loss of chromosome 22q in reduced dimensionality clusters. Cells from meningioma samples with loss of chromosome 22q and from dura samples with intact chromosome 22q were used for this analysis. **c**, Marker gene expression across reduced dimensionality clusters. **d**, Heatmap of differentially expressed genes across reduced dimensionality clusters, down sampled to 100 cells.

**Extended Data Fig 7. FOXM1 target genes and gene networks in meningioma.**

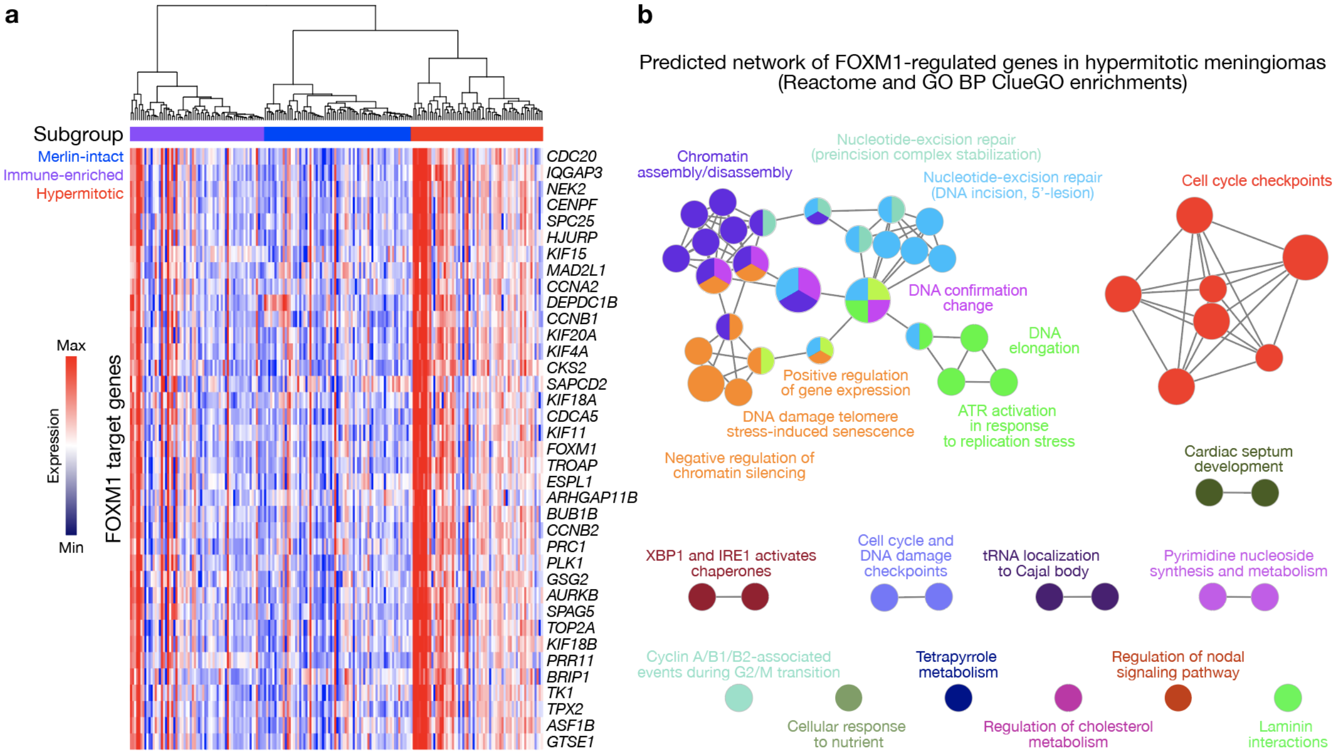

**Extended Data Fig. 7. FOXM1 target genes and gene networks in meningioma.**

**a**, Heatmap of relative FOXM1 target gene expression in meningiomas (n=200) across epigenetic groups.

**b**, Predicted network of FOXM1-regulated pathways in Hypermitotic meningiomas (see methods).

**Extended Data Fig. 8. The enhancer landscape across meningioma groups.**

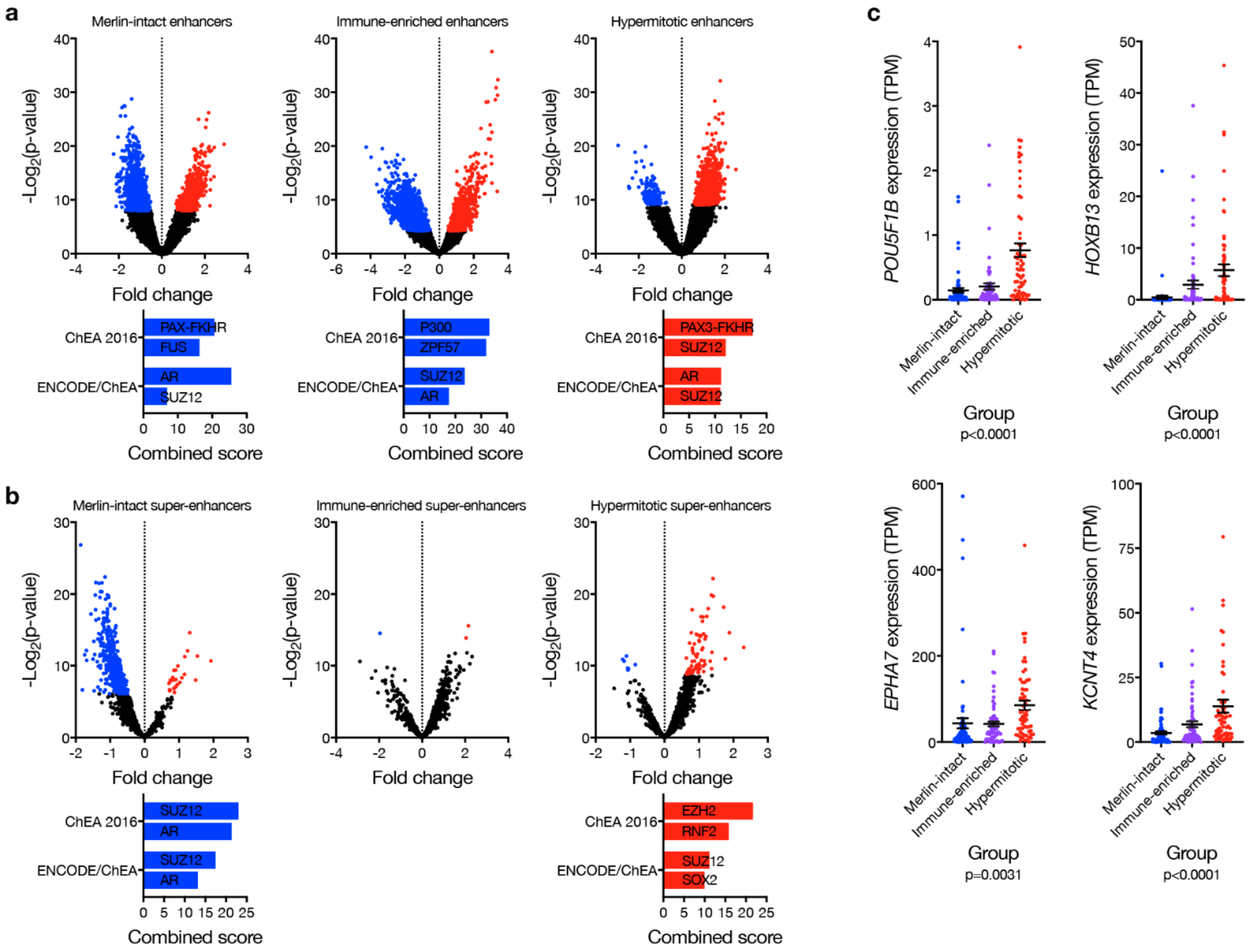

**Extended Data Fig. 8. The enhancer landscape across meningioma groups.**

**a**, Volcano plots of meningioma relative enhancer expression (n=25) across epigenetic groups (top), and gene ontology analyses (bottom), of differentially enriched (red) or suppressed (blue) enhancers. ChEA, ChIP-X Enrichment Analysis. **b**, Volcano plots of meningioma relative super-enhancer expression (n=25) across epigenetic groups (top), and gene ontology analyses (bottom), of differentially enriched (red) or suppressed (blue) super-enhancers. **c**, Meningioma transcripts per million (TPM) expression (n=200) of representative genes driving enhancer and super-enhancer ontologies from **a** and **b** across epigenetic groups (ANOVA).

### Extended Data Fig. 9. Prognostic models for meningioma recurrence.

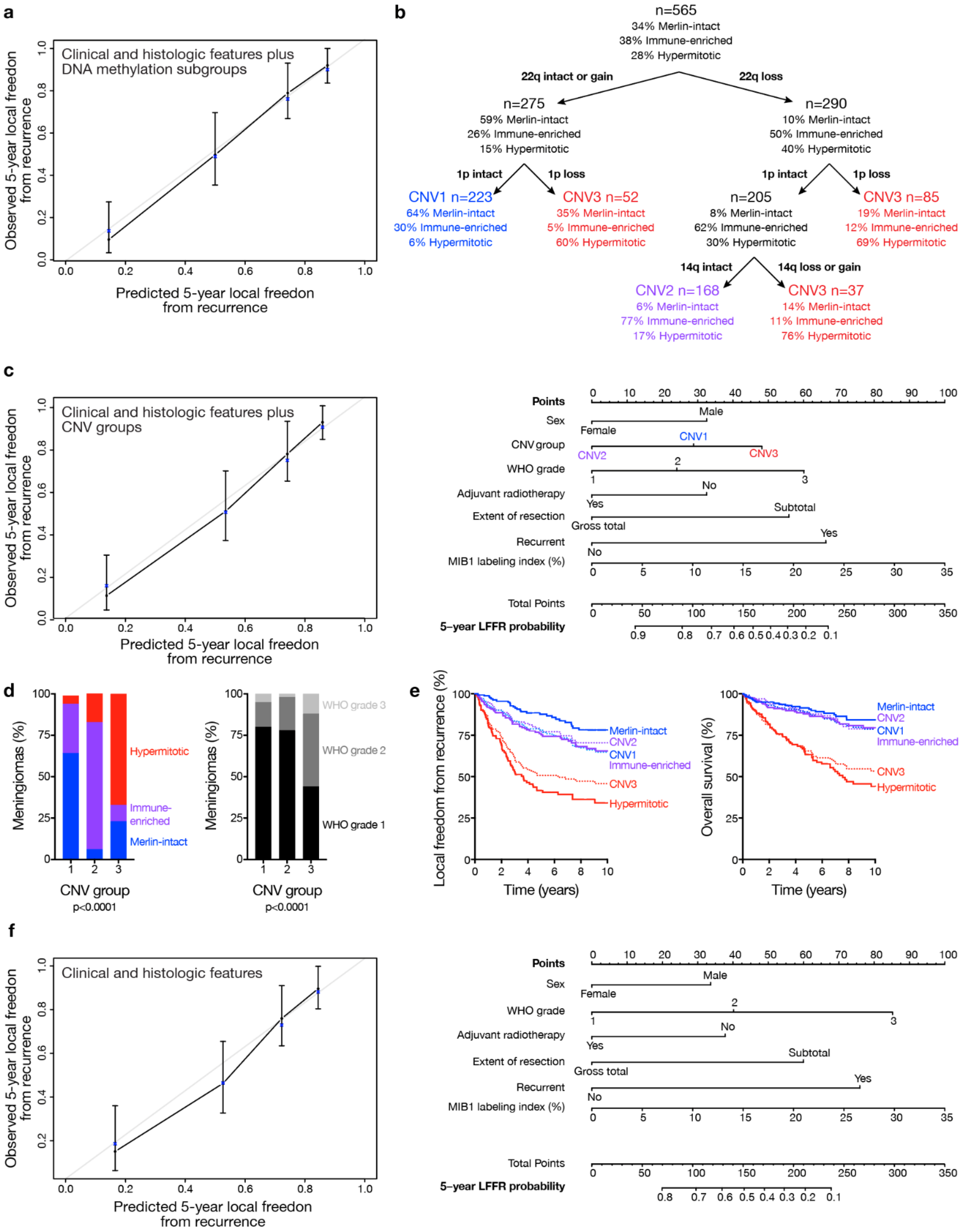

**Extended Data Fig. 9. Prognostic models for meningioma recurrence.**

**a**, Comparison of observed and predicted 5-year local freedom from recurrence (LFFR) from a model incorporating clinical, histologic, and epigenetic features (Fig. 5j). Blue asterisks on the calibration curve denote the bootstrap optimism-corrected estimated probabilities. **b**, Recursive partitioning analysis of meningiomas (n=565) by CNVs revealed 3 CNV-based groups. **c**, Comparison of observed and predicted 5-year LFFR from a model incorporating clinical, histologic, and CNV groups (left, n=201), used to generate a nomogram for meningioma LFFR (right, https://william-c-chen.shinyapps.io/RaleighLab_CNVSubgroupNomogram/). Each variable contributes points (top row) to the total score, which estimates the probably of 5-year LFFR (bottom 2 rows). **d**, Meningioma epigenetic groups and WHO grades (n=565) across CNV groups (Chi-squared tests). **e**, Kaplan-Meier curves for meningioma local freedom from recurrence and overall survival (n=565) comparing epigenetic and CNV groups. **f**, Comparison of observed and predicted 5-year LFFR from a model incorporating clinical and histologic features (left, n=201), used to generate a nomogram for meningioma LFFR (right, https://william-c-chen.shinyapps.io/RaleighLab_ClinicalVariablesNomogram/).

**Extended Data Table 1. Clinical features of the discovery and validation cohorts.**

**Extended Data Table 2. Univariable and multivariable regression outputs.**

**Extended Data Table 3. Meningioma *NF2* somatic short variants from targeted DNA sequencing across epigenetic groups.**

**Extended Data Table 4. Meningioma *TRAF7* somatic short variants and histologic subtypes associated with AKT1 E17K mutations across epigenetic groups.**

**Extended Data Table 5. Differentially expressed genes in $M10G^{dCas9\text{-}KRAB}$ cells.**

**Extended Data Table 6. Differentially expressed genes across meningioma epigenetic groups.**

**Extended Data Table 7. Differentially expressed genes in meningioma single-cell RNA sequencing clusters.**

**Extended Data Table 8. Transcription factor motif enrichment in differentially accessible enhancers and super-enhancers.**

### Methods

*Meningiomas and clinical data*

This study was approved by the UCSF Institutional Review Board (IRB #17-22324 and IRB #17-23196). Meningioma samples for the discovery cohort were selected from the UCSF Brain Tumor Center Biorepository and Pathology Core in 2017 to maximize the number of high-grade meningiomas and to acquire the longest clinical follow up possible for low-grade meningiomas. All WHO grade 2 and 3 meningiomas with available frozen tissue were included. For WHO grade 1 meningiomas, frozen samples in the tissue bank were cross-referenced for clinical follow-up data from a retrospective institutional meningioma clinical outcomes database (IRB #13-12587) to select all cases with available tissue and follow-up greater than 10 years (n=40). To achieve a discovery cohort of 200 cases, we also included additional frozen WHO grade 1 meningiomas with the longest follow-up available that was less than 10 years (n=47). The electronic medical record was reviewed for all patients in late 2018, and for patients treated prior to the advent of the electronic medical record, paper charts were reviewed those to achieve the maximal and most accurate clinical follow up. All available pathology material was reviewed for diagnostic accuracy by a board-certified neuropathologist (D.A.S.). WHO grading was performed according to contemporary criteria outlined in the most recent 4$^{th}$ edition of the WHO classification of tumors of the central nervous system[1]. Cases for which other mesenchymal central nervous system tumors (such as schwannoma or solitary fibrous tumor/hemangiopericytoma) remained in the differential diagnosis were excluded. The discovery cohort consisted of cases from 1991 to 2016. The validation cohort was a consecutive series of meningiomas from 2000 (the year of the most recent WHO update in meningioma classification) to 2019 that underwent resection and had frozen tissue available and clinical follow-up information from The University of Hong Kong. Meningioma recurrence was defined as new radiographic tumor on magnetic resonance imaging after gross total resection, or enlargement/progression/growth of residual tumor after subtotal resection.

*Nucleic acid extraction*

Frozen meningiomas were mechanically lysed using a TissueLyser II (QIAGEN) according to the manufacturer's instructions. DNA and RNA were extracted from lysed tissue using the AllPrep DNA/RNA/miRNA Universal Kit (#80224, QIAGEN). DNA and RNA quality were initially assessed using a NanoDrop One (Thermo Fisher Scientific). DNA samples with 260/280 values less than 1.8 or 260/230 values less than 1.6 were cleaned using ethanol precipitation and re-assessed. RNA samples with 260/280 values less than 1.8 or 260/230 values less than 1.6 were cleaned following the RNA Cleanup protocol within the RNeasy Mini Kit (#74106, QIAGEN). RNA samples were subsequently analyzed on a Bioanalyzer 2100 using the RNA 6000 Nano Kit (#5067-1511, Agilent Technologies). Only meningioma samples with both high-quality DNA (260/280 greater than 1.8 and 260/230 greater than 1.6) and high-quality RNA (RIN greater than 8) were used for DNA methylation profiling and RNA sequencing.

*DNA methylation profiling*

Genomic DNA was processed on the Illumina Methylation EPIC Beadchip (Illumina) according to manufacturer's instructions at the Molecular Genomics Core at the University of Southern California. Downstream analysis was performed in R version 3.5.3 with SeSAMe[2]. Probes were filtered and analyzed using the standard SeSAMe preprocessing pipeline. In brief, the following steps were performed: normal-exponential out-of-band background correction, nonlinear dye bias correction, p-value with out-of-band array hybridization masking, and β value calculation (β = methylated/[methylated+unmethylated]). Principal component analysis was performed in R using the base command ‘prcomp’ with the parameters ‘center = TRUE, scale. = FALSE’ on the β values. Variable probes were identified from the first three principal components. Specifically, the top 700 probes from each of those three principal components were selected (for a total of 2,100 probes), as ranked by the absolute value of their gene loading scores within their principal component. Duplicate probes were removed from the selected 2,100 probes and the probes with the lowest gene loading scores were culled until 2,000 variable probes remained. These 2,000 variable probes were used to perform unsupervised hierarchical clustering of the meningiomas (Pearson correlation distance, Ward’s method). K-means consensus clustering was performed to determine the optimal number of clusters using the ConsensusClusterPlus[3] R package, subsampling 1000 times per cluster number and using all 2,000 probes and 80% of samples per subsample.

A methylation profile-based multi-class support vector machine (SVM) classifier was generated using the caret R package. A linear kernel SVM was constructed using training data comprising 75% of randomly selected samples from the 200-sample discovery cohort with 10-fold cross validation. The 2,000 variable probes were used as variables. The remaining 25% of samples from the validation cohort were used to test the model and calculate its accuracy.

Leukocyte percentage within the tumor samples was calculated from DNA methylation using the ‘estimateLeukocyte’ command within the SeSAMe R package. In brief, the intensity of probes known to be uniquely hyper- or hypo-methylated in leukocytes were used to estimate the leukocyte percentage. Tumor purity of samples was estimated from DNA methylation using the PAMES[4] R package. In brief, we identified CpG sites and loci uniquely hyper- or hypo-methylated in tumor samples compared to 27 normal tissue controls and the intensity of hyper- or hypo-methylation at those sites were used to estimate tumor purity.

*DNA methylation-based copy number analysis*

CNV profiles from methylation data were generated as previously described[5] with the ‘cnSegmentation’ command within the SeSAMe R package, using the ‘EPIC.5.normal’ dataset from the sesameData package as a copy-number-normal control. Based on intensity value distributions, segments with mean intensity values less than -0.1 were identified as copy number loss, while segments with mean intensity values greater than 0.15 were identified as copy number gain. The percentage of the genome with copy number variation was determined by calculating the average number of segments per sample with mean intensity values less than -0.1 or greater than 0.15, weighted by segment length. CNV profiles by epigenetic group were generated by sampling the genome every 30,000 base pairs and determining the percentage of segments containing the sampled location that were identified as lost or gained. Specific gene loci were determined to be lost or gained if the entire locus was contained in a segment with mean intensity values less than -0.1 or greater than 0.15, respectively. CNV co-occurrence was visualized as an undirected weighted graph where nodes represented CNVs and edges represented the frequency of co-occurrence of CNV pairs. The size of each node was linearly proportional to the sum of total co-occurrences with other CNVs, while the thickness of each edge was linearly proportional to the frequency of the co-occurrence pair with a cutoff of frequency ≥ 5 for display. The igraph R package was used for graph visualization.

*RNA sequencing*

Library preparation was performed using the TruSeq RNA Library Prep Kit v2 (#RS-122-2001, Illumina) and 50 bp single-end reads were sequenced on an Illumina HiSeq 4000 to a mean of 42 million reads per sample at the IHG Genomics Core at the University of California San Francisco. Quality control of FASTQ files was performed with FASTQC[6]. Reads were trimmed with Trimmomatic[7] to remove Illumina adapter sequences, leading and trailing bases with quality scores below 20, and any bases that did not have an average quality score of 20 within a sliding window of 4 bases. Any reads shorter than 36 bases after trimming were removed. Reads were subsequently mapped to the human reference genome GRCh38[8] using HISAT2[9] version 2.1.0 with default parameters. For downstream expression analysis, we extracted exon level count data from the mapped HISAT2 output using featureCounts[10].

Differential expression analysis was performed in R version 3.5.3 with DESeq2[11], using the 'apeglm' parameter[12] to accurately calculate log fold changes and setting a false discovery rate of 0.05. Differentially expressed genes were identified as those with log fold changes greater than 1 and an adjusted p-value less than 0.05. Gene ontology analysis of differentially expressed genes was performed using Enrichr, and combined scores displayed are p-values combined with deviation z-scores[13,14]. Cell types within samples were deconvoluted by running xCell[15] with transcripts per million (TPM) values. In brief, the strength of gene expression patterns unique to different cell types were used to estimate the proportion of those cell type within each sample.

*TRAF7 variant calling from RNA sequencing*

Single nucleotide variants in *TRAF7* were identified from the RNA-seq data by following the Genome Analysis Toolkit's (GATK)[16] "RNAseq short variant discovery" Best Practices Workflow. In brief, the mapped HISAT output was processed by de-duplication and recalibration of base confidence scores. These processed reads were used an input for HaplotypeCaller[17] with the parameters '--dont-use-soft-clipped-bases true' and '-stand-call-conf 20' to identify somatic short variants (point mutations and small indels). The 'VariantFiltration' command within GATK was used to further filter the identified variants with the parameters '-window 35 -cluster 3 --filter-name "FS" -filter "FS > 30.0" --filter-name "QD" -filter "QD < 2.0".' Filtered variants were annotated using SnpEff[18,19].

*Targeted DNA sequencing*

A custom amplicon panel was designed with 100% coverage of all known coding exons of *NF2* (PGD364, Paragon Genomics). Genomic DNA was processed using this panel and the CleanPlex Target Enrichment and Library Preparation kit, following manufacturer's instructions (Paragon Genomics). Library quality was subsequently assessed on a TapeStation 4200 using the High Sensitivity D1000 Kit (#5067-5584, Agilent Technologies). Next, 150 bp paired-end reads were sequenced on an Illumina MiSeq v2 Micro at the Center for Advanced Technology at the University of California, San Francisco.

Quality control of FASTQ files was performed with FASTQC[6]. Reads were mapped to the *NF2* locus using Bowtie2[20]. Somatic short variants were identified using the Genome Analysis Toolkit (GATK)[16]. The mapped Bowtie2 output was processed with recalibration of base confidence scores. These processed reads were used an input for HaplotypeCaller[17] with the parameters '-ERC none' and '--max-reads-per-alignment-start 0' to identify somatic variants (point mutations and small indels). Somatic short variants were then filtered for a minimum total depth of 100 reads. Filtered variants were annotated using

SnpEff[18,19], and all but one *NF2* somatic short variant identified across groups were predicted to have 'HIGH' variant impact.

*Immunoblotting*

Immunoblot samples were prepared by lysis in radioimmunoprecipitation assay (RIPA) buffer containing Complete-Mini EDTA-free protease inhibitor (11836170001, Sigma-Aldrich) and PhosSTOP phosphatase inhibitor (04906837001, Sigma-Aldrich), followed by boiling in 6x Laemmli reducing buffer. Samples were separated on a 4-15% gradient TGX precast gel (4561086, Bio-Rad), and transferred onto nitrocellulose membranes (Whatman). Membranes were blocked in 5% TBST-milk, incubated in primary antibody, washed, and incubated in secondary antibodies (1:2000). Membranes were subjected to immunoblot analysis using Pierce ECL (Thermo Fischer Scientific) or SuperSignal West Femto (Thermo Fischer Scientific) substrates. Primary antibodies were used against Merlin (1:5000, ab88957, Abcam), GAPDH (1:10,000, MA515738, Thermo Fischer Scientific), Caspase 7 (1:1000, 9492, Cell Signaling Technology), IRF1 (1:1000, ab186384, Abcam), and IRF8 (1:1000, 5628S, Cell Signaling).

*Cell culture and organoids*

HEK293T, BenMen[21], IOMM-Lee[22], and CH157-MN[23] cells were cultured in Dulbecco's Modified Eagle Medium (DMEM) (#11960069, Life Technologies), supplemented with 10% fetal bovine serum (FBS) (#16141, Life Technologies), 1X GlutaMAX (#35050-061, Thermo Fischer Scientific), and 1X Penicillin/Streptomycin (#15140122, Life Technologies). DI-98 and DI-134 cells were generated as previously described[24], and cultured in Dulbecco's Modified Eagle Medium (DMEM) (#11960069, Life Technologies), supplemented with 7% fetal bovine serum (FBS) (#16141, Life Technologies), and 1X Penicillin/Streptomycin (#15140122, Life Technologies). M10G primary meningioma cells were generated as previously described[25], and cultured in a 1:1 ratio of DMEM/F12 media (#10565, Life Technologies) and Neurobasal media (#21103, Life Technologies), supplemented with 5% FBS (#16141, Life Technologies), B-27 supplement without vitamin A (#12587, Life Technologies), N-2 supplement (#17502, Life Technologies), 1X GlutaMAX (#35050, Life Technologies), 1mM NEAA (#11140, Life Technologies), 100U/mL Anti-Anti (#15240, Life Technologies), 20 ng/mL EGF (#AF-100-15, Peprotech), and 20 ng/mL FGF (#AF-100-18B, Peprotech). Human cerebral organoids were created from astrocytes induced from pluripotent human stem cells and co-cultured with meningioma cells as previously described[25].

Colorimetric proliferation assays were performed using the CellTiter 96 Non-Radioactive Cell Proliferation Assay (#G4100, Promega), according to manufacturer's instructions. For clonogenic assays, 150 cells were seeded in triplicate in 6 well plates. Cells were treated with either vehicle or drug both 1 and 6 days after seeding. After 10 total days of growth, cells were fixed in methanol for 30 minutes and stained with 0.01% crystal violet (C6158, Sigma-Aldrich) for 1 hour. Plates were rinsed with water three times, allowed to air dry, and imaged on a Zeiss Stemi 508 stereo microscope. Colony area was quantified by measuring total image intensity using ImageJ. Apoptosis assays were performed by treating cells with actinomycin D 0.5 µg/ml for 48 hours.

*CRISPRi cell line generation*

Lentiviral particles containing pMH0001 (UCOE-SFFV-dCas9-BFP-KRAB, #85969, Addgene)[26] were produced by transfecting HEK293T cells with standard packaging vectors using the *Trans*IT-Lenti

Transfection Reagent (MIR 6605, Mirus). M10G cells were stably transduced with these lentiviral particles to generate M10G$^{dCas9\text{-}KRAB}$ cells. Successfully transduced cells were isolated through selection of BFP positive cells using fluorescence activated cell sorting on a Sony SH800.

Single-guide RNA (sgRNA) protospacer sequences were individually cloned into the pCRISPRia-v2 vector (#84832, Addgene)[27], between the BstXI and BlpI sites, by ligation. Each vector was verified by Sanger sequencing of the protospacer. Protospacer sequences were sgNTC (GTGCACCCGGCTAGGACCGG), sg*NF2* (GGACTCCGCGCGCCTCTCAG), sg*USF1* (GAGATACCTAGGCCGGGAGA), sg*CDKN2A* (GTGGCCAGCCAGTCAGCCGA), sg*CDKN2B* (GACTCTGCCAGAGCGAGGCG). Lentivirus was generated (as described above) for each sgRNA expression vector. M10G$^{dCas9\text{-}KRAB}$ cells were independently transduced with lentivirus from each sgRNA expression vector, then selected to purity using 20 µg/mL puromycin over 7 days.

*shRNA gene suppression*

Lentiviral particles containing pLKO.1 shRNA targeting control (RHS6848, Dharmacon) or *NF2* (RHS3979-201768826 or RHS3979-201768830) were generated by transfecting HEK293T cells with standard packaging vectors (psPAX2 and pMD2.9) and shRNA plasmids using *Trans*IT-Lenti Transfection Reagent. After 48 hours of virus production, viral particles were sterilized through a 0.45 µM filter and added to meningioma cells with polybrene 10 µg/ml (TR-1003, MerkMillipore). A polyclonal population of shRNA positive cells was selected using puromycin 2 µg/ml.

*Immunofluorescence*

Immunofluorescence of primary meningioma cells was performed on glass coverslips. Cells were fixed in 4% paraformaldehyde, blocked in 2.5% BSA (Sigma) and 0.1% Triton X-100 (Sigma) in Phosphate Buffered Saline (PBS) for 30 min at room temperature (Thermo Fisher Scientific), and labeled with either anti-Ki67 (1:750, ab15580, Abcam) primary antibodies at room temperature for 1 h. Cells were labeled with Alexa Fluor secondary antibodies and either Hoechst 33342 (H3570, Thermo Fisher Scientific) or DAPI (D3571, Thermo Fisher Scientific) to mark DNA for 1 h at room temperature, and were mounted in ProLong Diamond Antifade Mountant (Thermo Fisher Scientific). For apoptosis assays, cells were washed in Annexin V binding buffer, stained with Annexin V (1:10, 550911, BD Bioscience) for 15 min, washed, labeled with DAPI to mark DNA for 1 h at room temperature, and mounted in ProLong Diamond Antifade Mountant.

Immunofluorescence of human meningiomas for lymphatic vessels was performed on 10 µm cryosections of frozen tissue embedded in OCT Compound (Thermo Fisher Scientific). Slides with tissue were fixed in cold acetone for 10 min, air dried, washed in PBS, permeabilized with 0.3% Triton-X 100 in PBS, and then washed again in PBS. Sections were blocked (2% BSA, 1% donkey serum, and 0.1% Triton-X 100 in PBS) for 30 min. Sections were then labeled with either anti-LYVE-1 (1:100, ab14917, Abcam) or anti-PROX-1 (1:100, AF2727, R&D Systems) primary antibodies at room temperature for 1 h. Slides were subsequently labeled with Alexa Fluor secondary antibodies and Hoechst 33342 to mark DNA for 1 h at room temperature, and were mounted in ProLong Diamond Antifade Mountant.

Dual immunofluorescence of human meningiomas for FOXM1 and Ki-67 was performed on 5 µm formalin-fixed, paraffin-embedded (FFPE) human meningioma sections. Following antigen retrieval using CC1 (950-124, Roche Diagnostics) for 32 min, sections were incubated and detected sequentially with rabbit monoclonal anti-Ki-67 (1:4, 30-9, Roche Diagnostics) and rabbit monoclonal anti-FOXM1 (1:600,

EPR17379, Abcam). Each primary antibody incubation was 32 min and single stained controls were used to verify specificity.

*Histology and immunohistochemistry*

Deparaffinization and rehydration of 5 µm FFPE human and mouse meningioma tissue sections and hematoxylin and eosin staining were conducted following standard procedures. Immunostaining was performed on an automated Ventana Discovery Ultra staining system. Immunostaining for Ki-67 was performed on 5µm FFPE murine meningioma sections using rabbit monoclonal anti-Ki-67 (1:6, 30-9, Roche Diagnostics) with primary antibody incubation for 16 min following CC1 antigen retrieval for 8 min.

CD3 immunoreactivity for each individual tumor was categorized qualitatively. Tumors were scored CD3 positive if multiple aggregates of CD3-positive lymphocytes were identified on examined sections, and were otherwise scored as CD3 negative. FOXM1 labeling index was quantified based on the total amount of nuclei with strong immunoreactivity for FOXM1 within a 200x field. Ki-67 labeling index was quantified based on the total amount of nuclei with strong immunoreactivity for Ki-67 within a 200x field. The labeling index for both FOXM1 and Ki-67 was averaged across two 200x fields for each individual tumor.

*Microscopy*

Fluorescence microscopy was performed on a Zeiss LSM 800 confocal laser scanning microscope with a PlanApo 20X air objective. Images were processed and quantified from representative regions of each tumor or coverslip using ImageJ.

Histologic and immunohistochemical features were evaluated using light microscopy performed using an Olympus BX43 microscope with standard objectives. Images were obtained and analyzed using the Olympus cellSens Standard Imaging Software package.

*Whole exome sequencing*

Exome capture and read sequencing were performed as previously described[28]. Paired-end sequence data were aligned using the Burrows-Wheeler Aligner to the reference human genome build hg19[29]. Duplicate removal, base quality recalibration, and multiple-sequence realignment were performed using Picard suite and Genome Analysis Toolkit[17,30]. Exome-based HLA Class I genotyping was performed using Polysolver and SOAP-HLA[31,32].

*Single Cell Isolation and Sequencing*

Fresh tissue samples were acquired from the operating room and transported to the laboratory space in PBS and on ice. Tissue samples were minced with sterile #10 scalpels (#4-410, Integra LifeSciences) then incubated at 37°C in a Collagenase Type 7 solution (#LS005332, Worthington) until digested (30-60 minutes). Collagenase was used at a concentration of 0.1 mg/mL for tumor and brain-tumor interface samples and at a concentration of 0.2 mg/mL for dura samples. Next, the samples were incubated in Trypsin-EDTA 0.25% (#25200056, Thermo Fisher Scientific) at 37°C for 5 minutes. The samples were incubated in 1X RBC lysis buffer (#00-4300-54, eBioscience) at room temperature for 10 minutes. Finally, samples were sequentially filtered through 70 µM and 40 µM cell strainers (#352350 and #352340, Corning) to generate a single-cell suspension.

Single cell suspensions were processed for single cell RNA-seq using onto a 10X Chromium controller, and libraries were generated using the Chromium Single Cell 3' Library & Gel Bead Kit v3 on a 10X Chromium controller (10X Genomics) using the manufacturer recommended default protocol and settings, at a target cell recovery of 5,000 cells per sample. Libraries were sequenced on an Illumina NovaSeq 6000, targeting >50,000 reads/cell, at the Center for Advanced Technology at the University of California, San Francisco.

*Single Cell Analysis*

Library demultiplexing, read alignment to the GRCh38 human reference genome, identification of empty droplets, and UMI quantification was performed using CellRanger version 3.0.2 (https://github.com/10XGenomics/cellranger). Cells with greater than 500 unique genes, less than 10,000 unique genes, and less than 20% of reads attributed to mitochondrial transcripts were retained. Single cell UMI count data were preprocessed in R version 3.6.1 with the Seurat[33,34] package version 3.0.1 using the sctransform[35] workflow, with scaling based on regression of UMI count and percentage of reads attributed to mitochondrial genes per cell. Dimensionality reduction was performed using principal component analysis and then principal components were corrected for batch effects using Harmony[36]. Uniform Manifold Approximation and Projection (UMAP) was performed on the reduction data with a minimum distance metric of 0.2 and Louvain clustering was performed using a resolution of 0.4. Marker identification and differential gene expression was performed in Seurat using a minimum fraction of detection of 0.75 and a minimum log-fold change of 0.5.

The presence or absence of somatic copy-number alterations (CNVs) in individual cells was assessed with CONICSmat[37]. Briefly, a two-component Gaussian mixture model was fit to the average expression values of genes on the chromosome 22q arm across all cells assessed. CNVs were assessed for cells from tumor samples with copy-number loss of the chromosome 22q arm at a bulk level as determined by DNA methylation and for cells from copy-neutral, normal dura samples. The command 'plotAll' from the CONICSmat R package was run with the parameters 'repetitions=100, postProb=0.75'. Cells with a posterior probability less than 0.15 were identified as tumor, while cells with a posterior probability greater than 0.85 were identified as normal. Clusters with greater than 80% of cells with an intact chromosome 22q arm were determined to be non-meningioma cell clusters. Standard immune, neural, and vascular markers found in the top 50 differentially expressed genes of the non-meningioma cell clusters were used to classify those clusters. Meningioma cell clusters were labeled based on gene programs and marker genes identified in the top 50 differentially expressed genes.

*Magnetic resonance imaging*

All patients in the discovery cohort underwent preoperative magnetic resonance imaging (MRI) on clinical scanners at either 1.5 or 3.0 Tesla field strength. MRI protocols varied across the study period, but all patients included for image analysis had T1 pre- and post-intravenous gadolinium-based contrast agent administration sequences, T2-weighted spin echo sequences, and T2-weighted fluid attenuated inversion recovery (FLAIR) sequences. Post-contrast T1 images evaluated in this study were high-resolution 3D, allowing for multiplanar reconstruction. Evaluation of meningioma proximity to dural venous sinuses was performed qualitatively by a board-certified radiologist with a Certificate of Added Qualification in Neuroradiology (J.E.V-M.) on post-contrast T1 images. Meningiomas were classified as involving a dural venous sinus if they abutted a dural reflection or frankly invaded the sinus.

*ChIP sequencing and enhancer/super-enhancer analysis*

H3K27ac ChIP sequencing data were derived from 25 previously reported meningiomas overlapping with our discovery cohort[24]. Enhancer and super enhancer analyses were performed as previously described[24]. Briefly, FASTQ reads were trimmed to remove low quality reads and adaptors with TrimGalore and uniquely mapped reads were aligned to the human reference genome hg19/GRCh37 with the Burrows-Wheeler Aligner[29]. SAMtools was used to sort and index BAMs, and PCR duplicates were removed with PicardTools. Peaks were called using MACS2 with the default log2 fold change enrichment of 2 compared to input and a p-value cutoff of $10^{-5}$. Consensus peaksets and normalized H3K27ac densities were generating using the DiffBinds R package. Peaks present in at least 2 tumor samples were used to generate a consensus peakset and overlapping peaks were merged. Peaks on chromosome X or Y and peaks intersecting ENCODE blacklisted regions v1 on haplotype chromosomes were excluded from analysis. Bigwig tracks were generated using DeepTools (v3.1.2) with RPKM normalization and were visualized using Integrative Genomics Viewer software. Super-enhancers were called using ROSE with default parameters. Gene set enrichment networks were generated using ClueGO and visualized in Cytoscape. Prediction of FOXM1-regulated genes was performed by first identifying FOXM1 binding motif sites using Homer to scan across the genome for the known FOXM1 motif. These sites were then intersected with H3K27ac peaks in the consensus meningioma peakset. These were annotated to the nearest gene using Homer. This gene set was intersected with genes positively and significantly (FDR<0.05) correlated with FOXM1 expression and genes upregulated in the Hypermitotic subgroup versus others (FDR<0.05)

*ChIP quantitative reverse transcriptase PCR*

ChIP qPCR was performed using the EZ-Magna ChIP A/G Chromatin Immunoprecipitation Kit (Millipore), according to manufacturer's instructions. Briefly, cells were fixed in 1% formaldehyde and sonicated to fragment sizes of 200-800 bp. Samples were incubated overnight with 10 µg USF1 antibody (ab180717, Abcam) or IgG antibody bound to protein A and protein G magnetic beads. After antibody incubation, samples were washed once each with high salt, low salt, lithium chloride and TE buffers. Samples were de-crosslinked by incubation at 65°C for 4 hours, followed by incubation at 95°C for 10 minutes, and then purified using a PCR purification kit (Invitrogen). qPCR was performed using PowerUp SYBR Green Master Mix (#A25918, Thermo Fisher Scientific). QPCR primer sequences targeting gene promoters were NC1-F (5'-AAAAGCAGCCCATCTCTGTG-3'), NC1-R (5'-TGGGAGACAGAGCAAGACTC-3'), NC2-F (5'-TTCTAACTTGGCTCGGGCATC-3'), NC2-R (5'-TCGCCTAACCTCTTCAGCTTC-3'), CDK6-F (5'-TTGTCTTTCGGCTCGCTGTC-3'), CDK6-R (5'-AATCCTCAGGCCCAGAAAGG-3'), FOXM1-F (5'-CACCGGAGCTTTCAGTTTGTTC-3'), and FOXM1-R (5'-TTCCGTCACGTGACCTTAACG-3')

*RNA extraction, cDNA synthesis, and quantitative reverse transcriptase PCR*

RNA was extracted from cultured cells using the RNeasy Mini Kit (#74106, QIAGEN) according to manufacturer's instructions. cDNA was synthesized from extracted RNA using the iScript cDNA Synthesis Kit (#1708891, Bio-Rad). Real-time qPCR was performed using PowerUp SYBR Green Master Mix (#A25918, Thermo Fisher Scientific) on a QuantStudio 6 Flex Real Time PCR system (Life Technologies). Real-time QPCR primer sequences were GAPDH-F (5'-GTCTCCTCTGACTTCAACAGCG-3'), GAPDH-R (5'-ACCACCCTGTTGCTGTAGCCAA-3'), CDKN2A-F

(5’-ATGGAGCCTTCGGCTGACT-3’), CDKN2A-R (5’-GTAACTATTCGGTGCGTTGGG-3’), CDKN2B-F (5’-ACGGAGTCAACCGTTTCGGGAG-3’), CDKN2B-R (5’-GGTCGGGTGAGAGTGGCAGG-3’), USF1-F (5’-CTGCTGTTGTTACTACCCAGG-3’), USF1-R (5’-TCTGACTTCGGGGAATAAGGG-3’), CDK6-F (5’-TCTTCATTCACACCGAGTAGTGC-3’), CDK6-R (5’-TGAGGTTAGAGCCATCTGGAAA-3’). Real-time qPCR data were analyzed using the ΔΔCt method relative to *GAPDH* expression.

*Mice*

Patient-derived xenograft experiments were performed by implanting 3 million CH-157MN cells into the flank of 5-6-week-old NU/NU mice (Harlan Sprague Dawley Inc.). Animals in the treatment arm were gavaged with 100 μg/g abemaciclib in 0.5% methylcellulose vehicle daily starting 12 days after injection, until protocol-defined endpoints. For Kaplan-Meier survival analysis, events were recorded when tumors reached the protocol-defined size of 2000 $mm^3$.

*Patients*

Patients were treated with Abemaciclib 100 mg *per os* twice daily. Treatment was held in the setting myelosuppression (absolute neutrophil count less than 1.5), and diarrhea was managed with over the counter medications. Meningioma volumes on serial magnetic resonance imaging studies were determined using MIM (MIM Software Inc).

*Meningioma nomograms*

Prognostic models for LFFR were generated using multivariable Cox regression via the survival R package. The proportional hazards assumption was confirmed by visual inspection of the Schoenfeld residuals and the Schoenfeld global test[38]. Variables included in the final model were selected by a two-step process, first by a univariable Cox regression threshold of p≤0.05, followed by selection of features with greatest variable importance as estimated by the Breiman permutation method using concordance as the model metric[39]. The top 7 features were selected to allow for at least 10 events per variable in the final model. This process was repeated for creation of the methylation group model and CNV group model. Models with and without the candidate biologic group variable were compared using the bootstrapped time-dependent delta-AUC and delta-Brier-score for LFFR at 5 years[40]. The survAUC R package was used to calculate time-dependent AUC and Brier-scores. Nomograms based on the final Cox models were visualized using the ‘nomogram’ function of the rms R package. Cox model calibration of 5-year LFFR was estimated using the ‘calibrate’ function of the rms R package with default settings, utilizing Kaplan-Meier estimates, bootstrapping, and with an average group size of 50 subjects per calibration level. Unless otherwise specified, all bootstrap procedures were performed with 500 iterations. Recursive partitioning analysis of CNV and methylation groups was performed using the rpart R package, with a minimum of 30 observations per split attempt and minimum of 15 observations per terminal leaf. The optimal complexity parameter was determined by 5-fold cross-validation, with selection of the most parsimonious model defined as the model with fewest splits and no more than one standard-error above the error of the best model[41]. Finally, interactive web-based nomogram graphical user interfaces were created using the DynNom R package.

**Extended references**


1. Louis, D., Ohgaki, H., Wiestler, O. & Cavenee, W. *WHO Classification of Tumours of the Central Nervous System*. (2016).
2. Zhou, W., Triche, T. J., Laird, P. W. & Shen, H. SeSAMe: reducing artifactual detection of DNA methylation by Infinium BeadChips in genomic deletions. *Nucleic Acids Res.* **46**, e123–e123 (2018).
3. Wilkerson, M. D. & Hayes, D. N. ConsensusClusterPlus: a class discovery tool with confidence assessments and item tracking. *Bioinformatics* **26**, 1572–1573 (2010).
4. Benelli, M., Romagnoli, D. & Demichelis, F. Tumor purity quantification by clonal DNA methylation signatures. *Bioinforma. Oxf. Engl.* **34**, 1642–1649 (2018).
5. Capper, D. *et al.* DNA methylation-based classification of central nervous system tumours. *Nature* **555**, 469–474 (2018).
6. Andrews, S. *FastQC*.
7. Bolger, A. M., Lohse, M. & Usadel, B. Trimmomatic: a flexible trimmer for Illumina sequence data. *Bioinforma. Oxf. Engl.* **30**, 2114–2120 (2014).
8. Schneider, V. A. *et al.* Evaluation of GRCh38 and de novo haploid genome assemblies demonstrates the enduring quality of the reference assembly. *Genome Res.* **27**, 849–864 (2017).
9. Kim, D., Paggi, J. M., Park, C., Bennett, C. & Salzberg, S. L. Graph-based genome alignment and genotyping with HISAT2 and HISAT-genotype. *Nat. Biotechnol.* **37**, 907–915 (2019).
10. Liao, Y., Smyth, G. K. & Shi, W. featureCounts: an efficient general purpose program for assigning sequence reads to genomic features. *Bioinforma. Oxf. Engl.* **30**, 923–930 (2014).
11. Love, M. I., Huber, W. & Anders, S. Moderated estimation of fold change and dispersion for RNA-seq data with DESeq2. *Genome Biol.* **15**, 550 (2014).
12. Zhu, A., Ibrahim, J. G. & Love, M. I. Heavy-tailed prior distributions for sequence count data: removing the noise and preserving large differences. *Bioinformatics* **35**, 2084–2092 (2019).
13. Chen, E. Y. *et al.* Enrichr: interactive and collaborative HTML5 gene list enrichment analysis tool. *BMC Bioinformatics* **14**, 128 (2013).
14. Kuleshov, M. V. *et al.* Enrichr: a comprehensive gene set enrichment analysis web server 2016 update. *Nucleic Acids Res.* **44**, W90-97 (2016).
15. Aran, D., Hu, Z. & Butte, A. J. xCell: digitally portraying the tissue cellular heterogeneity landscape. *Genome Biol.* **18**, 220 (2017).
16. Auwera, G. A. V. der *et al.* From FastQ Data to High-Confidence Variant Calls: The Genome Analysis Toolkit Best Practices Pipeline. *Curr. Protoc. Bioinforma.* **43**, 11.10.1-11.10.33 (2013).
17. DePristo, M. A. *et al.* A framework for variation discovery and genotyping using next-generation DNA sequencing data. *Nat. Genet.* **43**, 491–498 (2011).
18. Cingolani, P. *et al.* A program for annotating and predicting the effects of single nucleotide polymorphisms, SnpEff: SNPs in the genome of Drosophila melanogaster strain w1118; iso-2; iso-3. *Fly (Austin)* **6**, 80–92 (2012).
19. Cingolani, P. *et al.* Using Drosophila melanogaster as a Model for Genotoxic Chemical Mutational Studies with a New Program, SnpSift. *Front. Genet.* **3**, 35 (2012).
20. Langmead, B. & Salzberg, S. L. Fast gapped-read alignment with Bowtie 2. *Nat. Methods* **9**, 357–359 (2012).
21. Püttmann, S. *et al.* Establishment of a benign meningioma cell line by hTERT-mediated immortalization. *Lab. Invest.* **85**, 1163 (2005).

22. Lee, W. H. Characterization of a newly established malignant meningioma cell line of the human brain: IOMM-Lee. *Neurosurgery* **27**, 389–395; discussion 396 (1990).
23. Tsai, J.-C., Goldman, C. K. & Gillespie, G. Y. Vascular endothelial growth factor in human glioma cell lines: induced secretion by EGF, PDGF-BB, and bFGF. *J. Neurosurg.* **82**, 864–873 (1995).
24. Prager, B. C. *et al.* The Meningioma Enhancer Landscape Delineates Novel Subgroups and Drives Druggable Dependencies. *Cancer Discov.* (2020) doi:10.1158/2159-8290.CD-20-0160.
25. Magill, S. T. *et al.* Multiplatform genomic profiling and magnetic resonance imaging identify mechanisms underlying intratumor heterogeneity in meningioma. *Nat. Commun.* **11**, 4803 (2020).
26. Adamson, B. *et al.* A Multiplexed Single-Cell CRISPR Screening Platform Enables Systematic Dissection of the Unfolded Protein Response. *Cell* **167**, 1867-1882.e21 (2016).
27. Horlbeck, M. A. *et al.* Compact and highly active next-generation libraries for CRISPR-mediated gene repression and activation. *eLife* **5**, e19760 (2016).
28. Vasudevan, H. N. *et al.* Comprehensive Molecular Profiling Identifies FOXM1 as a Key Transcription Factor for Meningioma Proliferation. *Cell Rep.* **22**, 3672–3683 (2018).
29. Li, H. & Durbin, R. Fast and accurate short read alignment with Burrows-Wheeler transform. *Bioinforma. Oxf. Engl.* **25**, 1754–1760 (2009).
30. McKenna, A. *et al.* The Genome Analysis Toolkit: a MapReduce framework for analyzing next-generation DNA sequencing data. *Genome Res.* **20**, 1297–1303 (2010).
31. Shukla, S. A. *et al.* Comprehensive analysis of cancer-associated somatic mutations in class I HLA genes. *Nat. Biotechnol.* **33**, 1152–1158 (2015).
32. Cao, H. *et al.* An integrated tool to study MHC region: accurate SNV detection and HLA genes typing in human MHC region using targeted high-throughput sequencing. *PloS One* **8**, e69388 (2013).
33. Butler, A., Hoffman, P., Smibert, P., Papalexi, E. & Satija, R. Integrating single-cell transcriptomic data across different conditions, technologies, and species. *Nat. Biotechnol.* **36**, 411–420 (2018).
34. Stuart, T. *et al.* Comprehensive Integration of Single-Cell Data. *Cell* **177**, 1888-1902.e21 (2019).
35. Hafemeister, C. & Satija, R. Normalization and variance stabilization of single-cell RNA-seq data using regularized negative binomial regression. *Genome Biol.* **20**, 296 (2019).
36. Korsunsky, I. *et al.* Fast, sensitive and accurate integration of single-cell data with Harmony. *Nat. Methods* **16**, 1289–1296 (2019).
37. Müller, S., Cho, A., Liu, S. J., Lim, D. A. & Diaz, A. CONICS integrates scRNA-seq with DNA sequencing to map gene expression to tumor sub-clones. *Bioinforma. Oxf. Engl.* **34**, 3217–3219 (2018).
38. Schoenfeld, D. Partial residuals for the proportional hazards regression model. *Biometrika* **69**, 239–241 (1982).
39. Breiman, L. Random Forests. *Mach. Learn.* **45**, 5–32 (2001).
40. Song, X. & Zhou, X.-H. A SEMIPARAMETRIC APPROACH FOR THE COVARIATE SPECIFIC ROC CURVE WITH SURVIVAL OUTCOME. *Stat. Sin.* **18**, 947–965 (2008).
41. Hastie, T., Tibshirani, R. & Friedman, J. *The Elements of Statistical Learning: Data Mining, Inference, and Prediction*. (2009).
